# Integrating container cleaning practices into a Zambian Community Health Club program - evidence from a pilot study

**DOI:** 10.1101/2025.10.29.25339087

**Authors:** Tracy Zhang, Yanwen Li, Nathan Ndonde, Elijah Mutafya, Gabrielle String

## Abstract

Communities relying on drinking-water storage routinely document containers as a reservoir for pathogens when they are not properly cleaned. Although lab-based research provides guidance on cleaning protocols, gaps remain in understanding: 1) what methods are practiced, and 2) method effectiveness at reducing biofilm growth. In partnership with AFMAC, this pilot explored biofilm contamination in household water storage containers of four Zambian communities engaged in WASH-promoting community health clubs and health promoters. A mixed-methods design was used: 15–20 households per community completed a knowledge, attitudes, and practices survey and provided whole-container biofilm samples. Four focus group discussions with program members and four interviews with AFMAC staff explored container cleaning sensitizations. Findings revealed household awareness of container cleaning, inconsistent cleaning practices, and high levels of biofilm in 100% of storage containers. The absence of a standardized container cleaning module, reliance on visual cues as a proxy for microbial safety, and disproportionate burden placed on women underscore critical gaps in current programming and highlight container cleaning as a critical pathway to interrupt recontamination. This pilot highlights the need for locally adapted, practical protocols using accessible materials. Advancing container hygiene through integrated, adaptive, and scalable approaches is essential for reducing public health risks from unsafe water storage and improving point-of-use water quality.

**Synopsis:** Field evidence on household water storage container cleaning is limited. The study found high biofilm levels and inconsistent cleaning of household water storage containers, revealing a gap between knowledge and practice in maintaining safe water quality.

**Table of Content Graphic:** 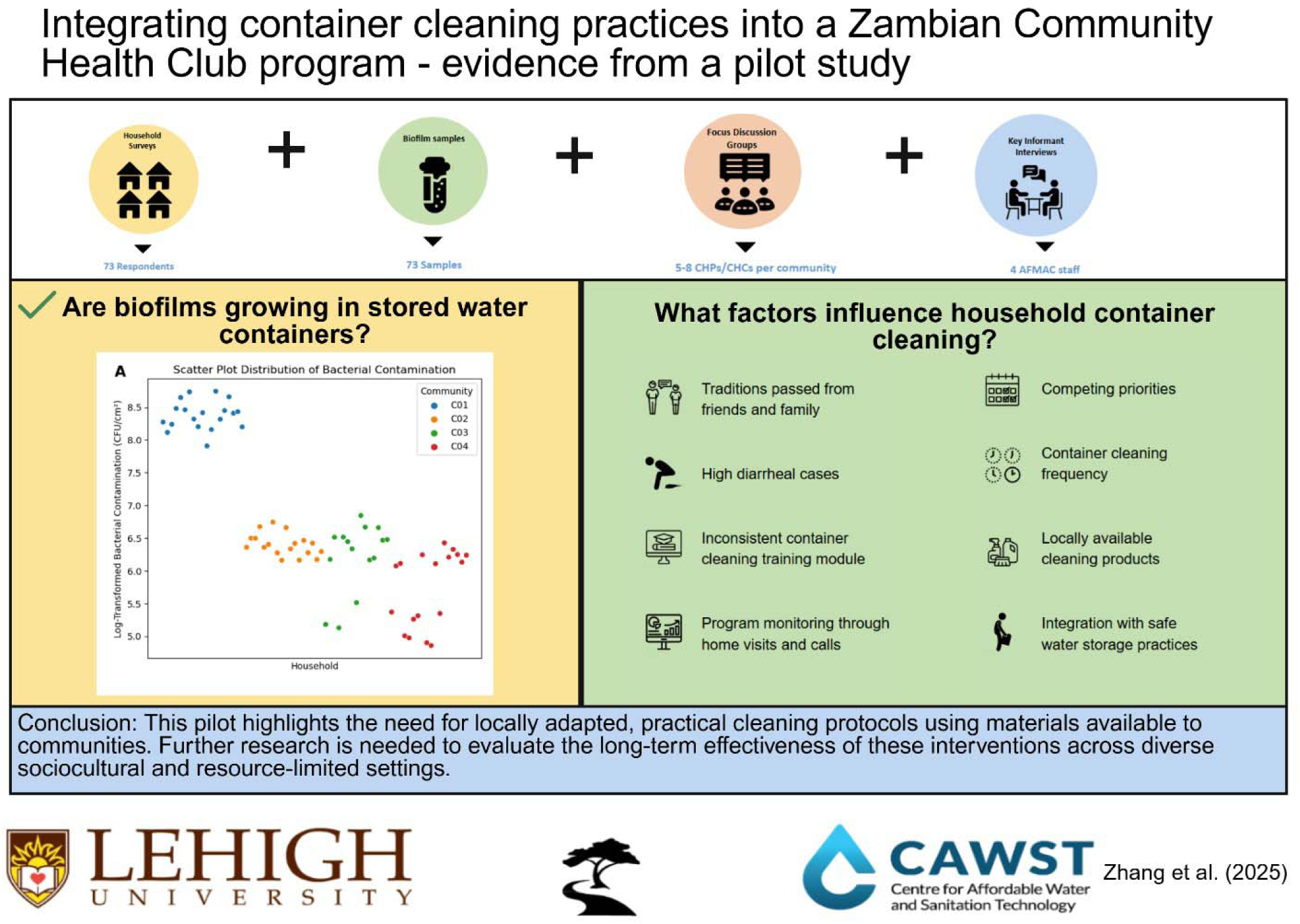

## 1.0 INTRODUCTION

Approximately 74% of the global population has access to safely managed water services that are free from contamination, available when needed, and accessible on-premise^1^. Despite this progress, many households still rely on collection and storage of drinking water, introducing risks of microbiological recontamination^2,3^. Safe water storage (SWS) practices—such as using opaque containers with narrow openings, drawing water via taps or pouring, and regularly cleaning with soap—are recommended to minimize recontamination and support 2030 UN Sustainable Development Goal targets 6.1 and 3.9^4–7^. These targets aim to achieve universal and equitable access to safe and affordable drinking water and reduce the number of deaths and illnesses from hazardous environmental contaminations, respectively.

Microbial risks from storage arise in part due to biofilms, which are communities of microorganisms that colonize surfaces like pipes and container interiors^8^ (Supplemental Background S1). These biofilms can act as persistent microbial reservoirs, increase chlorine demand in stored water and contribute to the deterioration of drinking water quality^9–11^. While laboratory studies reported chlorine-based disinfectants are effective at reducing bacterial contamination^8^, they are not always accessible or affordable in LMICs. Other materials, such as brushes, sand, and rocks, have been used for container cleaning but their use and effectiveness differ for container types. For instance, a field study in Uganda found that using sand in jerricans reduced *E. coli* but not total coliform levels, while another study found brushes to reduce total coliform and E.coli for containers with wide openings and a spigot^12–14^ (Background S1). Overall, the field evidence on household water storage container cleaning is limited, and laboratory-tested methods do not guarantee best practices in varied real-world settings.

Cleaning frequency is also critical to maintaining safe water quality. One study found that chlorine disinfection combined with weekly container cleaning helped reduce recontamination during 24-hour storage, but the optimal cleaning frequency remains unclear^11^. A separate cross-sectional study found most households clean containers prior to refilling, often every 3 days, yet water stored beyond 3 days without cleaning has been associated with increased health risks^15,16^. Experimental work suggests biofilm development can occur within 24 hours when using untreated water and within 7–14 days following treatment via silver-impregnated filters^17^.

Zambia, like many LMICs, lacks national policy guidance on household water treatment and storage (HWTS)^18^. Rural areas continue to face challenges with unimproved water sources (28.2% use) and sanitation access (47.98% unimproved)^19^. While a randomized trial showed improved microbiological water quality and reduced diarrhea when households used both filtration and safe storage^20,21^, few studies have explored household container cleaning specifically in the Zambian context.

To address this gap, we investigated water storage container cleaning practices in Zambian households with the Africa MANZI Center (AFMAC), a local NGO promoting water, sanitation, and hygiene (WASH). AFMAC implements community-led health interventions via Community Health Promoters (CHPs) and Community Health Clubs (CHCs), supported by the Centre for Affordable Water and Sanitation Technology (CAWST)^22–24^. CHP is a volunteer-led program engaging individually with households while CHC is a self-sustaining group of community members adapted from Zimbabwe’s Africa Ahead model (Background S4). These programs provide WASH education, distribute resources, and guide behavior change through locally adapted lesson plans^25^.

## 2.0 MATERIALS AND METHODS

### 2.1 Study Design and Setting

We designed a mixed-methods study to examine household container biofilm contamination and cleaning practices among four AFMAC partner communities. The study included key informant interviews (KII) with AFMAC staff, focus group discussions (FGD) with CHP and CHC members, household surveys, and household drinking water and container biofilm sampling. The study protocol was approved by the Lehigh University Institutional Review Board (Protocol# 2137976-1) and the ERES Converge Institutional Review Board in Zambia (Reference# 2024-Feb-015). Staff familiar with the communities and Bemba language were hired as research assistants (RA) and trained in ethics, informed consent, and study tools.

Data collection occurred in four randomly selected communities where AFMAC implemented community health initiatives; Kaloko, Mwange, and George Compound had both CHP and CHC programs while Chibili Village had only a CHC program. Top community health concerns as noted by local health centers are malaria, diarrhea, and respiratory tract infection (Background S5).

### 2.2 KIIs and FGDs

KIIs were conducted over Zoom with four current or former AFMAC staff involved in the implementation of CHP and CHC programs. The semi-structured interviews focused on understanding program design and implementation; organization of container cleaning sensitizations; and, opinions on how they have affected household container cleaning practices (Methods S1.2). Each interview was conducted in English, audio-recorded with an auto-generated transcript reviewed for accuracy. FGDs were completed with 5-8 CHPs and CHC members from each community. FGDs were held with men and women together, as CHP and CHC programs are mixed gender. RAs used a discussion guide focused on CHC and CHP program activities; container cleaning lesson plans and materials; community behavior and practices; and, opinions regarding container cleaning sensitizations (Methods S1.3). Discussions were conducted in Bemba, audio-recorded, and digitally transcribed and translated to English by RAs.

### 2.3 Household Survey and Sampling

Households were surveyed randomly, with every Nth household chosen for inclusion (Methods S1.1) until 15-20 households were enrolled per community. Given time and resource constraints, RAs reached fewer households in some communities compared to others. Verbal consent was obtained from each household before administering the survey to an adult over age 18 (preferably the female head of household). The household survey consisted of 59 questions and observations on household demographics; knowledge, attitudes, and practices towards drinking water storage container cleaning; and, physicochemical water quality sampling (Methods S1.4). The survey was written in English and deployed through mWater survey application (mWater, New York, NY) on the RAs’ phones. During the survey, RAs verbally translated questions to Bemba at the comfort of the respondent. Additionally, they took photos of the inside and outside of sampled drinking water storage containers and measured the sizes of the containers to approximate the surface area biofilm could grow on. RAs asked respondents, “May I have a cup of drinking water?” and used this sample to measure physicochemical water parameters on-site, including temperature, pH, electrical conductivity (EC), and total dissolved solids (TDS) using an Apera PC60 Premium multiparameter portable probe (Apera, Columbus, OH).

At the conclusion of the household survey, RAs asked participants to temporarily pour their stored water into a holding container. Then, RAs used a sterile long-handled scrub brush and 200 mL of sterile water to scrub the inside surfaces of the household’s drinking water container to dislodge matter into the sterile water. The sample was collected in a sterile WhirlPak® bag with sodium thiosulfate (Nasco, Fort Atkinson, WI), stored on ice, and transported to AFMAC’s field-established laboratory for processing within 12 hours. Because of the risk that dislodged microbial matter might remain in the sampled container, 1 capful of 2.25% chlorine solution was added to the container, shaken to coat surfaces, and allowed to sit in contact for at least 30 minutes before participants emptied, rinsed, and poured their drinking water back into the container.

### 2.4 Biofilm Processing

Heterotrophs encompass a wide variety of microorganisms, including bacteria, yeasts, and molds; heterotrophic plate count (HPC) is used as an indicator of general microbial content of water to assess the cleanliness of water distribution systems^26^, especially when temperature increases to 37 °C trigger regrowth. Biofilm samples were diluted appropriately using sterile buffer water, vacuum filtered aseptically through a 0.45-micron filter (Fisherbrand, Hampton, NH), plated on m- HPC agar (NEOGEN, Lansing, MI) for 48 hours at 35°C for detection of bacteria, and enumerated with 10% duplicates and 5% blanks. Colonies were counted and concentrations calculated by averaging plate counts within a countable range (10-200 CFU/plate) accounting for dilution factors. Concentrations were divided by the surface area of the container (CFU/cm^2^).

### 2.5 Data Analysis

Data was securely uploaded by researchers to Box Webpage (Box, Redwood City, CA) and analyzed using MAXQDA (VERBI Software, Berlin, Germany). An inductive coding approach was applied to KII and FGD analysis, whereby codes emerged and were assigned labels through close reading without predefined themes^27^. Emergent codes were grouped into broader themes and interpreted in relation to the research question. Codebooks were generated and intercoder reliability ensured consistency and validity across the codes identified^27^.

Broader KII themes included: motivation to implement WASH programs, differences between CHP and CHC programs, and AFMAC assessment of programs’ impact (Figure S1). Broader FGD themes included: influence of community engagement, tools used to promote behavior change, cleaning methods, cultural factors, and persistent challenges in sustaining cleaning routines (Figure S2).

Data from household surveys, on-site water quality testing and microbiological testing were recorded on the mWater application (mWater, New York, NY). Results were exported as a .csv file into Microsoft Excel 2010 (Microsoft, Redmond, WA) and cleaned. Descriptive statistics and statistical analysis were completed using Python (Python Software Foundation, Wilmington, DE). Non-parametric statistical tests, including Kruskal-Wallis and Mann-Whitney-U, were conducted to determine significant SWS behaviors related to biofilm. Chi-square tests with Monte Carlo simulation were run to determine differences by community in SWS behaviors. For all statistical tests, p<0.05 were considered statistically significant.

## 3.0 RESULTS

This study was conducted in four communities in Ndola, Zambia, from April to May 2024. We conducted four KIIs with AFMAC staff, four FGDs with CHPs and CHCs, surveyed 73 households, and processed 73 biofilm samples. These efforts provided an understanding of WASH integrated activities in community health programs (Results S1), including program implementation and monitoring efforts; how CHCs and CHPs engage with communities; households’ knowledge and safe storage practices; and how container cleaning relates to biofilm contamination and water quality (Table S2, Results S3).

### 3.1 Program Implementation and Monitoring

AFMAC staff and FGD participants highlighted a need for container cleaning lessons within CHC and CHP training as their program motivation. Community members’ knowledge of container cleaning came from traditions passed from friends and family. These practices included not cleaning containers, rinsing the container, or cleaning only when visibly dirty. Households also lacked the necessary material needed for adequate container cleaning, such as soap, brushes, and chlorine. A KII participant shared, “Even if I don’t put sand, even if I don’t put anything, it’s okay, because my parents, my grandmother, my grandfather used to do that. ‘We never used to die’… that’s what they believe in.” AFMAC staff also noticed high diarrheal cases in the community, especially during the rainy season. As shared by AFMAC staff, “…There used to be too many diarrhea cases. And every rainy season, there used to be diarrhea cases”.

The CHC and CHP programs had separate training modules resulting in inconsistent container cleaning lessons. CHC training modules, adapted from Zimbabwe Africa Ahead, focused on SWS topics with container cleaning briefly mentioned. Elaborating on the CHP training, one interviewee explained that AFMAC worked with CAWST to create their own locally adapted training for CHPs with formal container cleaning lessons. One interviewee, discussing training module differences, shared, “when you’re doing a bio-sand filter training, you also talk about the storage container…. But maybe the in-depth details of the cleaning weren’t covered as much before. Of course, you talk about making sure that you clean it with some soap or some form of paste or detergent,” overall noting, “CHPs have a formal container cleaning training…So, it was piloted with the CHPs, not the CHCs.”

AFMAC staff monitor program impact through home visits and phone calls with CHC facilitators and promoters, stressing the importance of frequent communication. To monitor their work, CHPs submit monthly forms reporting activities and AFMAC staff attend CHC meetings. Furthermore, one interviewee shared that they were able to see a decrease in disease incidence before and after intervention implementation, as their protocol is to, “work with the environmental health technicians. Before we do any intervention, we go and find out the rate of diseases that are in that community.”

AFMAC staff and program members observed behavior change and a reduction in waterborne diseases following sensitization efforts. One FGD participant noted, “We used to have a lot of diarrhea cases, but they have reduced. Most people never cared about their hands, water, or latrines, but after our continuous sensitization, people have improved”. AFMAC staff and program members believe interactive lessons improved understanding and adoption of container cleaning practices. AFMAC staff noted, “… But now, under the training in this workshop, we emphasize cleaning the container every day before refilling it. So, you’d find that every time you visit the household, you see a clean container, unlike before when they’d wait maybe a week until there’s visible dirt outside.”

### 3.2 CHP and CHC Engagement with the Community

Both CHP and CHC programs engaged local leaders to build trust and credibility within their communities, employing different community engagement methods to foster relationships and active participation. CHCs foster peer accountability and shared responsibility, with members monitoring each other’s progress on WASH tasks thereby creating a supportive environment for members to share challenges and solutions. Whereas, CHPs focus on personalized household visits, providing tailored advice and monitoring practices at an individual level. The peer-centered approach allows facilitators to lead by example, as noted “When they see that others in the community are no longer getting sick, they want to know what they are doing differently”. Furthermore, AFMAC staff shared, “Being part of the club makes them feel like they are learning and doing something valuable”.

#### 3.2.1 Engaging with Lessons

Both programs conduct home visits to identify areas for WASH improvement, employing teaching tools including demonstrations and visual aids paired with conversation (Results S2).

Lesson cards illustrating correct cleaning practices help bridge literacy gaps. One FGD participant stated, “The pictures show exactly what to do, so even those who can’t read understand”.

Facilitators demonstrate step-by-step cleaning techniques, such as scrubbing with sand or ash and disinfecting with chlorine (Results S2). These interactive methods increase community buy-in and knowledge retention.

Furthermore, participants noted, “Usually we give them examples and consequences of not cleaning their storage containers” referring to health risks from microorganisms. AFMAC staff emphasized container cleaning is a collective responsibility that can further the spread of disease in the community. They shared, “We used to tell them to say there are microbes that you cannot see with your own eyes…And because that happens, then it is important we ensure where we are storing our water, it is thoroughly cleaned and this is not only for you, but also for the entire community. So, if one of the community members’ household has cases of diarrhea, dysentery or typhoid, meaning they are also affected…”

#### 3.2.2 Challenges in Sustained Behavior and Practice

While many households responded positively to sensitization efforts, long term behavior change remains a challenge. A participant shared, “We do notice some change at first but as days goes, they begin to revert back to their usual way of doing things”. Program members have found that peer accountability and regular check-ins helped reinforce behavior, explaining, “When we return for follow-ups, they are reminded of what they learned and try to do better”.

Program members repeatedly stressed the need for additional resources to support sensitization efforts such as sponges, brushes, chlorine, and lesson cards. FGD participants noticed the lack of cleaning materials hindered consistency and program impact. A FGD participant said, “They might want to clean the container, but they don’t have soap or anything to clean with, so they skip it”. A FGD participant shared, “If we had more demonstration containers and cleaning tools, we could teach better”. Mobility was another concern, with participants requesting bicycles to reach remote households. “Some homes are far, and we can’t visit them often without transport,” one facilitator explained. Uniforms or badges were also suggested to enhance credibility and recognition during household visits.

#### 3.2.3 Promoted Container Cleaning and SWS Methods

Research participants shared locally adapted approaches for effective container cleaning taught to communities. One FGD participant explained, “Some people use ashes when they don’t have soap. It works well to remove dirt and disinfect”. Another FGD participant added, “After cleaning the container with sand and soap, we tell them to disinfect with chlorine or, if they don’t have that, boil water to rinse it”. Furthermore, participants shared varied recommendations for container cleaning frequency from before every refill to after a specified number of days to prevent biofilm buildup. One FGD participant shared, “But now, under the training in this workshop, we emphasize cleaning the container every day before refilling it.”, while another shared, “…After cleaning the container, I told her that this is how a storage container should be cleaned. I advised her to be doing this every 2 days.”

Container cleaning sensitizations were emphasized to be a holistic WASH practice along with SWS practices. One FGD participant explained, “We educate them that treating water without cleaning the container is pointless because germs in the container will make the water dirty again”. Container cleaning is integrated into broader lessons on water treatment and hygiene as demonstrated by, “We tell them to store clean water in separate containers, one for drinking and another for other uses, so they don’t mix and risk contamination”.

### 3.3 Household Characteristics and SWS Container Biofilm Growth

Most respondents were men (75%); 62% of respondents were 25-49 years old (Table S1). A majority of respondents attended school (88%). There were 80% female-headed households, and 94% of heads of households could read and write. Households averaged 5.4 family members. Of the 28 households that reported at least one diarrhea case in the last 7 days, an average of 1.5 members per household had diarrhea.

Top reported drinking water sources were protected (49%) and unprotected (27%) dug wells, tube well/borehole (11%), public tap/standpipe (5%), and surface water (4%). Households spent on average 10 minutes with a maximum of 30 minutes collecting and returning with water. Households mainly used the same container to collect and store their drinking water (65%). More than half (57%) of the households had refilled their storage container the day of the survey (Table 1). Seven households (9%) indicated they had treated their water, using sand filters or liquid chlorine, the day of the survey (57%) or the day prior (28%).

**Table 1.** Household Demographics, Water Handling, and Container Practices. Data from household surveys (n=73) is presented alongside the mean biofilm log_10_HPC CFU/cm^2^ for the corresponding household survey category. P-value by community assesses differences in participant responses by community. P-value by biofilm assesses differences in biofilm growth by participant response.

| Variables | Households n=73<br>(percent) |  | p-value<sup>b</sup> | Biofilm<sup>a</sup><br>Mean (range) |  | p-value<sup>c</sup> |
| --- | --- | --- | --- | --- | --- | --- |
| N (%) Households included in the study |  |  |  |  |  |  |
| Community 1 | 19 | (27) | – | 8.4 | (7.9-8.7) | < 0.001*** |
| Community 2 | 18 | (26) |  | 6.4 | (6.2-6.7) |  |
| Community 3 | 15 | (21) |  | 6.2 | (5.1-6.8) |  |
| Community 4 | 18 | (26) |  | 5.7 | (4.9-6.4) |  |
| N (%) Top reported drinking water sources |  |  |  |  |  |  |
| Protected dug well | 36 | (49) | <0.01** | 6.6 | (4.9-8.7) | < 0.001*** |
| Unprotected dug well | 20 | (27) |  | 6.1 | (5.0-6.7) |  |
| Tubewell / borehole | 8 | (11) |  | 7.6 | (5.5-8.7) |  |
| Public tap / standpipe | 5 | (5.5) |  | 8.4 | (8.2-8.7) |  |
| N (%) Respondents store water in collection container | 48 | (66) | <0.05* | 6.6 | (4.9-8.7) | 0.054 |
| N (%) Last time container was filled |  |  |  |  |  |  |
| Today | 42 | (58) | 0.129 | 6.8 | (5.0-8.7) | 0.37 |
| Yesterday | 25 | (34) |  | 6.5 | (4.9-8.5) |  |
| Several days ago | 6 | (8.2) |  | 6.9 | (6.2-8.2) |  |
| N (%) Household treated water | 7 | (9.6) | <0.01** | 6.9 | (6.4-8.7) | 0.13 |
| N (%) Water retrieval method |  |  |  |  |  |  |
| Dips cup | 61 | (84) | 0.21 | 6.7 | (4.9-8.7) | 0.84 |
| Pour water out of container | 9 | (12) |  | 7 | (5.0-8.7) |  |
| From container’s tap | 3 | (4) |  | 6.6 | (6.4-6.8) |  |
| N (%) Top types of containers in use |  |  |  |  |  |  |
| Bucket | 65 | (89) | 0.46 | 6.7 | (4.9-8.7) | 0.58 |
| Jerrycan | 8 | (11) |  | 7.1 | (5.0-8.7) |  |
| N (%) Top reported reasons for their choice of storage containers |  |  |  |  |  |  |
| Convenient | 49 | (67) | 0.67 | 6.6 | (4.9-8.7) | 0.66 |
| Low cost / affordable | 26 | (36) |  | 7.1 | (4.9-8.7) |  |
| Easy to use | 26 | (36) |  | 6.7 | (4.9-8.7) |  |
| Sturdy | 11 | (15) |  | 6.7 | (5.3-8.6) |  |
| N (%) Container opening size |  |  |  |  |  |  |
| Very wide | 36 | (49) | - | 6.6 | (4.9-8.7) | - |
| Wide | 29 | (40) |  | 6.8 | (5.0-8.6) |  |
| Mean (range) volume of the container (L) | 19 | (2.5- 60) | - | - | - | - |
| N (%) Location of the storage container |  |  |  |  |  |  |
| Inside, on ground | 49 | (67) | 0.26 | 6.7 | (5.0-8.7) | 0.16 |
| Inside, raised on table | 16 | (22) |  | 6.4 | (4.9-8.2) |  |
| On top of another bucket | 5 | (6.8) |  | 6.8 | (4.9-8.3) |  |
| N (%) Observed exterior of container was visibly dirty | 29 | (40) | <0.05* | 7.2 | (5.1-8.7) | <0.05* |
| N (%) Observed interior of container was visibly dirty | 13 | (18) | 0.43 | 7 | (6.1-8.4) | 0.25 |
| N (%) Observed interior and exterior container combined |  |  |  |  |  |  |
| Both interior and exterior of the container are NOT visibly dirty | 40 | (55) | 0.08 | 6.4 | (4.9-8.7) | <0.05* |
| Exterior is visibly dirty, but interior is not | 20 | (27) |  | 7.3 | (5.1-8.7) |  |
| Both interior and exterior of the container are visibly dirty | 9 | (12) |  | 7 | (6.1-8.4) |  |
| Interior is visibly dirty, but exterior is not | 4 | (5.5) |  | 7 | (6.5-8.4) |  |
| N (%) Observed interior dirtiness level |  |  |  |  |  |  |
| Moderately dirty | 27 | (40) | <0.05* | 6.8 | (5.2-8.7) | 0.15 |
| Lightly used | 22 | (30) |  | 6.2 | (4.9-8.7) |  |
| Very dirty | 16 | (22) |  | 7.1 | (5.0-8.7) |  |
| N (%) Observed condition of the container interior |  |  |  |  |  |  |
| White particles/sand | 21 | (29) | 0.99 | 6.3 | (5.3-8.4) | 0.1 |
| Black/brown particles | 5 | (6.8) |  | 7.1 | (5.4-8.4) |  |
| Green or black algal growth | 4 | (5.5) |  | 6.3 | (6.1-6.5) |  |
| N (%) Household cleans their container | 64 | (88) | 0.99 | 6.7 | (4.9-8.7) | 0.47 |
| N (%) Household last cleaned the container, n=65 |  |  |  |  |  |  |
| Today | 36 | (55) | 0.1 | 6.8 | (5.0-8.7) | 0.69 |
| Yesterday | 23 | (35) |  | 6.6 | (4.9-8.4) |  |
| Several days ago | 6 | (9.2) |  | 6.9 | (6.2-8.2) |  |
| N (%) Clean the container at home, n=65 | 64 | (99) | 1 | 6.7 | (4.9-8.7) | 0.22 |
| N (%) Materials used to clean the container, n=65 |  |  |  |  |  |  |
| Water | 61 | (94) | 0.38 | 6.7 | (4.9-8.7) | 0.1 |
| Sponge/brush | 53 | (82) |  | 6.6 | (4.9-8.7) |  |
| Soap | 34 | (52) |  | 6.6 | (4.9-8.7) |  |

<sup>a</sup> Three households had plate growth that exceeded the maximum detection limit and were excluded from mean HPC concentrations (n=70)
<sup>b</sup> P-value assessing significant differences between the communities
<sup>c</sup> P-value assessing significant difference between biofilm contamination level
\* p-value <0.05; \*\* p-value <0.01; \*\*\* p-value <0.001

Biofilm contamination across all communities significantly varied 4.9-8.7 log_10_CFU/cm^2^ (p<0.01) (Table 1, Figure S3). Biofilm growth was more variable in Community 4 but clustered in the upper range for Community 1 (Figure 1, Figure S4). Water sources also differed by community and biofilm contamination (p<0.01). Public taps/standpipes, found only in Community 1, had the highest average biofilm contamination, while containers storing water from unprotected dug wells, found in Communities 2 and 4, had the lowest average contamination. Using separate collection and storage containers differed by community (p<0.05) but not by biofilm contamination (p=0.054). Communities 3 and 1 had the highest percentage of households, (62.5%, 40%), that used different containers for water collection and storage. Water treatment behaviors differed by community (p<0.01); most households that treated their collected water were from Community 3 (Table 1). Additionally, 84% of households retrieved water by dipping cups. Water retrieval methods did not differ statistically by community (p=0.21) or biofilm contamination (p=0.84).

**Figure 1.**
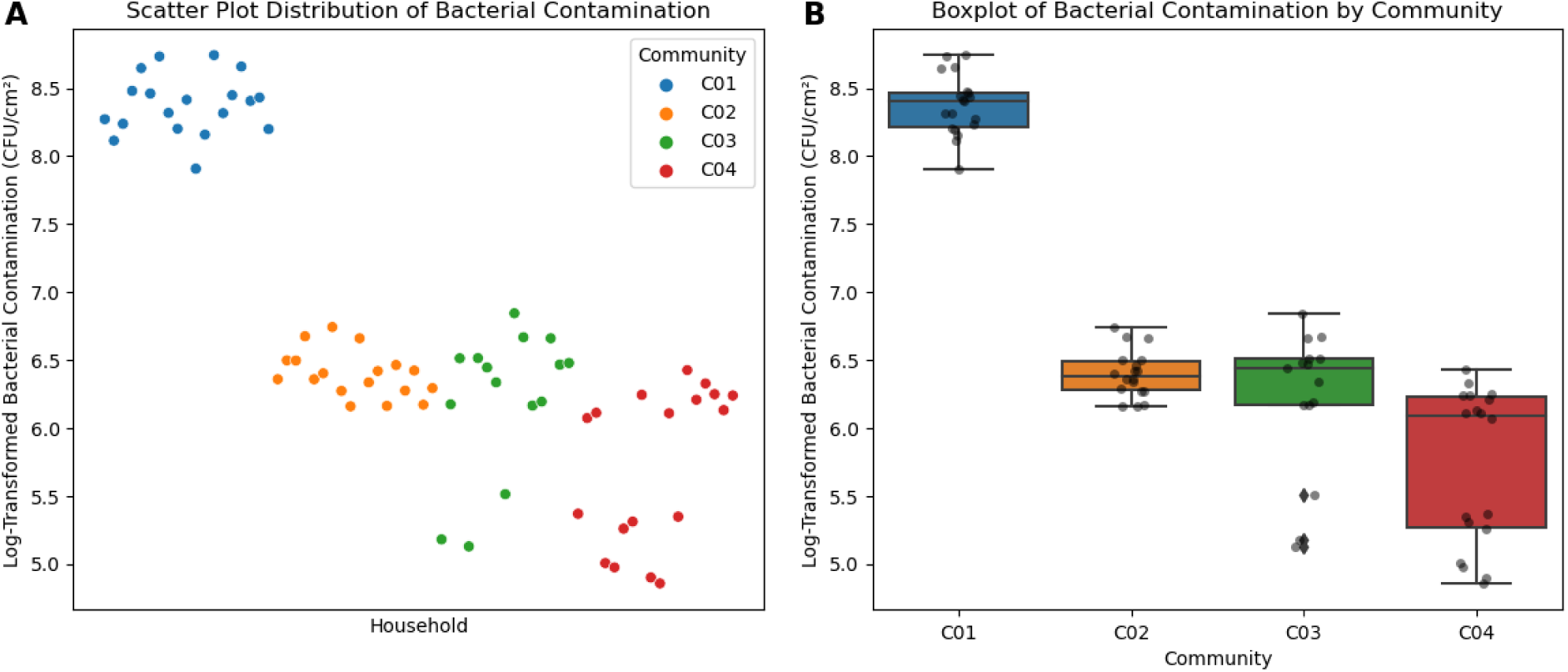
Bacterial Contamination of Household Containers: Individual and Community Patterns. - Panel A depicts the clustering and variability of bacterial contamination across all households. Each data point represents a household, and the color represents their associated community. Panel B depicts a box plot of bacterial contamination separated by community. Each box plot contains the 25th percentile, median, and 75th percentile of each group’s distribution of values as represented by the horizontal line respectively. The vertical lines extend to 1.5x the interquartile range, and each black dot represents an individual household.

Top reported container types were buckets with wide openings (89%) and jerrycans with small openings (11%). Containers were selected for convenience (67%), affordability (35%), ease (35%), sturdiness (15%), and limited options (4%). Containers averaged 18.8 L volume, and were stored inside either on the ground (67%) or raised on the table (21%). RAs observed 39% of container exteriors were visibly dirty and 1% were cracked; interiors were moderately dirty (40%), lightly used (30%), or very dirty (21%). Container interiors had white sand particles (28%), brown/black particles (6%), and green or black algal growth (5%) upon observation (Table 1). Most households (87%) cleaned their containers on the survey day (55%) or the day before (35%), at home (98%). For container cleaning, households used a mixture of water, sponge/brush, soap, detergent, rocks and sand, ash and chlorinated water. Top reported materials were water (93%), sponge/brush (81%) and soap (52%) (Table 1); the top reported combinations were “water, sponge or brush, and soap” and “water and sponge or brush” (Figure 2).

**Figure 2.**
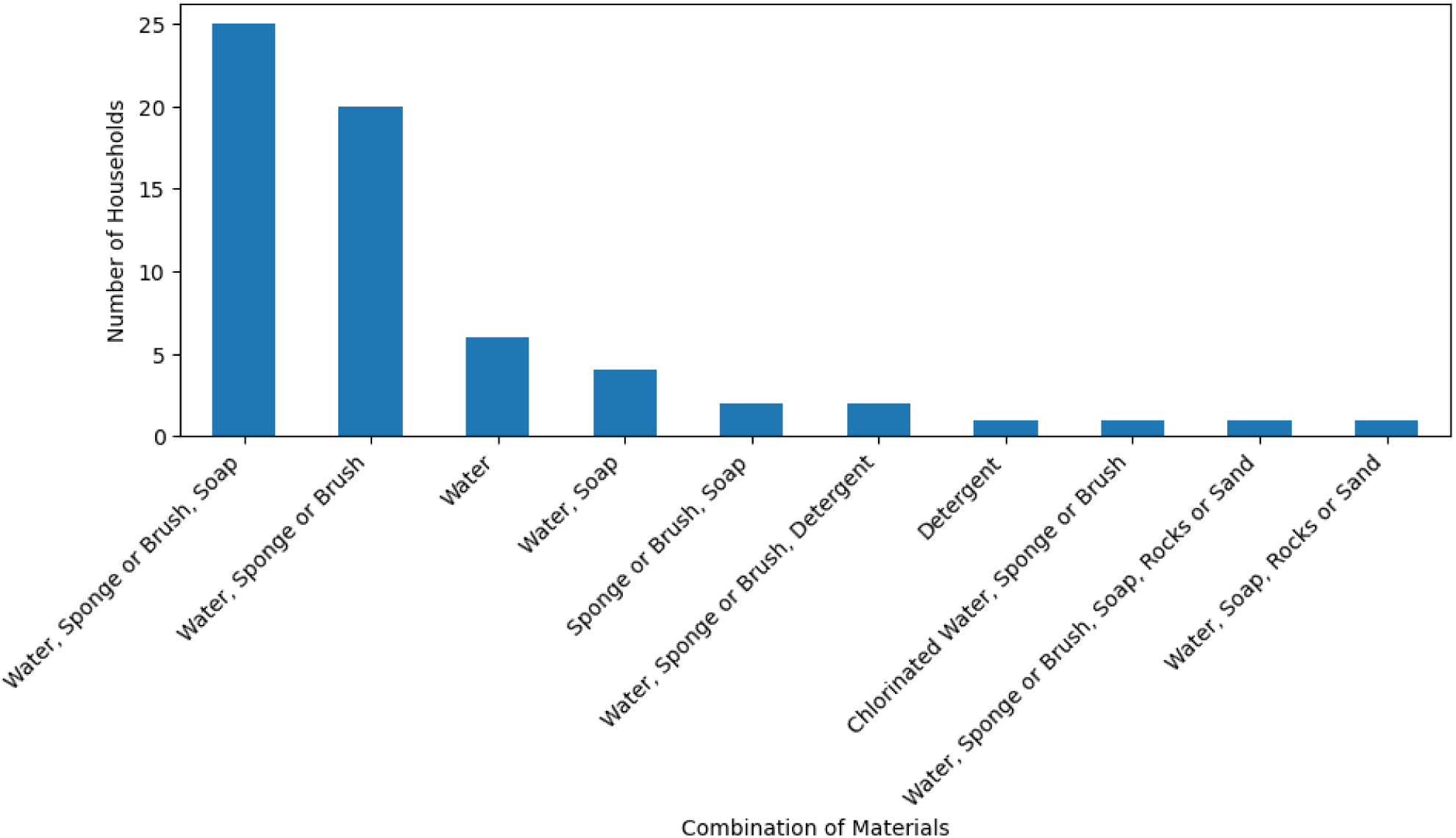
Combination of Container Cleaning Materials by Household. - Bar plot of the combination of materials utilized by households to clean their containers.

Observations of the container exterior being visibly dirty differed by community (p<0.05), but not by biofilm contamination (p=0.15) (Table 1). Containers with visibly dirty exteriors (P<0.05) had an average biofilm contamination of 7.2 log_10_CFU/cm^2^, while those without had an average of 6.4 log_10_CFU/cm^2^. Community 1 had the highest average households (65%) with observed dirty exteriors, and Community 4 had the lowest (16.7%). Further analysis found significant differences in biofilm contamination (p<0.05) across categories of interior and exterior visible dirtiness; however, pairwise comparisons suggest there were no significant findings on which combinations of interior and exterior dirtiness differed significantly from one another (Table 1). Lastly, container interior and exterior appearance significantly differed by enumerator (p=0.03), indicating there may have been bias due to the enumerators’ visual assessments of ‘dirtiness.’

Most respondents (97%) believed containers should be cleaned with the majority cleaning daily (89%). Most respondents (97%) also believed they could get sick from unclean containers; top reported illnesses include diarrhea (93%), cholera (74%), and stomachache (66%) (Table 2). The top reported reasons respondents knew when to clean their container were: container exterior is dirty (64%), container interior is dirty (61%), water is not clear (27%), based on the last time it was cleaned (16%), and before refilling the container (9%) (Table 2). Respondents believed water was unsafe to drink when it looked dirty (97%) or tasted/smelled bad (41%). Few respondents (10%) had been formally taught to clean water storage containers from workshops and friends/family. The five respondents who attended workshops averaged 11 sessions delivered by AFMAC, CHPs, or CHCs.

**Table 2:** Knowledge and Attitudes of Drinking Water Storage and Container Cleaning. Data from household surveys (n=73) is presented alongside the mean biofilm log_10_ HPC CFU/cm^2^ for the corresponding household survey category. P-value by community assesses differences in participant responses by community. P-value by biofilm assess differences in biofilm growth by participant response.

| Variables | Households<br>n=73<br>(percent) |  | p-value<sup>b</sup> | Biofilm<sup>a</sup><br>Mean (range) |  | p-value<sup>c</sup> |
| --- | --- | --- | --- | --- | --- | --- |
| N(%) Respondents believe containers should be cleaned | 73 | (100) | - | 6.7 | (4.9-8.7) | - |
| N(%) Frequency respondents believe containers should be cleaned |  |  |  |  |  |  |
| Daily | 65 | (89) | 0.06 | 6.7 | (4.9-8.7) | 0.36 |
| Weekly | 8 | (11) |  | 7.4 | (6.2-8.7) |  |
| N(%) Respondents believe they can get sick from unclean containers | 71 | (97) | 1 | 6.7 | (4.9-8.7) | 0.22 |
| N(%) Top reported sicknesses that you can get from unclean containers, n=71 |  |  |  |  |  |  |
| Diarrhea | 66 | (93) | 0.68 | 6.7 | (4.9-8.7) | 0.43 |
| Cholera | 53 | (75) |  | 6.7 | (5.0-8.7) |  |
| Stomachache | 47 | (66) |  | 6.6 | (4.9-8.7) |  |
| N(%) Top reported reasons respondents know when to clean their container |  |  |  |  |  |  |
| Outside of container is dirty | 47 | (64) | <0.01** | 6.7 | (4.9-8.7) | 0.19 |
| Inside of container is dirty | 45 | (62) |  | 6.7 | (4.9-8.7) |  |
| Water is not clear | 20 | (27) |  | 6.8 | (5.1-8.7) |  |
| N(%) Top reported reasons that respondents believe water is not safe to drink |  |  |  |  |  |  |
| Water looks dirty | 71 | (97) | 0.71 | 6.7 | (4.9-8.7) | 0.71 |
| Water tastes/smells bad | 30 | (41) |  | 6.5 | (4.9-8.7) |  |
| Water is not treated | 3 | (4.1) |  | 6.7 | (5.3-8.3) |  |
| N(%) Respondent has NOT been taught to clean water storage containers | 65 | (89) | 0.59 | 6.8 | (4.9-8.3) | 0.9 |
| N(%) Respondent's source of container cleaning knowledge, n=8 |  |  |  |  |  |  |
| Workshops/sensitizations | 5 | (63) | 0.57 | 6.5 | (4.9-8.3) | 0.71 |
| Friends and family | 4 | (50) |  | 6.1 | (4.9-6.7) |  |
| Mean (range) number of AFMAC container cleaning workshop, n=5 | 11.8 | (1-18) | - | - | - | - |

<sup>a</sup> Three households had plate growth that exceeded the maximum detection limit and were excluded from mean HPC concentrations (n=70)
<sup>b</sup> P-value assessing significant differences between the communities
<sup>c</sup> P-value assessing significant difference between biofilm contamination level
\* p-value <0.05; \*\* p-value <0.01; \*\*\* p-value <0.001

Reported indicators for cleaning containers differed by community (p<0.05). Average bacterial contamination (6.7 log_10_CFU/cm^2^) was similar whether households reported using exterior or interior dirtiness as a cleaning indicator (Table 2). Community 1 had the highest percentage of households that cleaned their containers based on an unspecified time since the last cleaning (p=<0.0001). Although containers from households with daily cleaning beliefs (6.7 log_10_CFU/cm^2^) had lower average biofilm contamination than those with weekly cleaning beliefs (7.4 log_10_CFU/cm2), the difference was not significant (p=0.36) (Table 2). Respondents taught to clean containers had lower average biofilm contamination, especially those taught by their friends and family, but this difference was not significant (p=0.9) (Table 2).

## 4.0 DISCUSSION

This pilot study highlights three key findings: 1) biofilm contamination in stored drinking water containers was high; 2) the disconnect between household cleaning practices and container contamination; and 3) strategies to integrate the promotion of SWS and container cleaning into ongoing community health programs (Discussion S1). These results call for the broad need for more tailored and specific recommendations on container cleaning to address this drinking water recontamination pathway.

### 4.1 Biofilm contamination of stored drinking water containers was high

In our study, biofilm contamination among household containers was high, ranging 4.9 -8.7 log_10_CFU/cm^2^. No standards currently exist for interpreting HPC biofilm levels and associated drinking water^26,28^; however, 100-500 CFU/mL or non-noticeable changes in the concentration of HPC is recommended^26^. Budeli et al., found HPC levels of inoculated coupons increased over 21 days from 2.3 to 5.76 CFU/cm² in untreated water and 20.2 to 227.8 CFU/cm² in water filtered through bio-sand filters^17^.

Interestingly, we observed a significant difference in average biofilm contamination between communities, from 5.7 log_10_CFU/cm^2^ (Community 4) to 8.4 log_10_CFU/cm^2^ (Community 1), potentially due to differences in water source. Containers storing water from public tap/standpipe, only used in Community 1, had the highest average biofilm contamination (8.4 log_10_CFU/cm^2^). In contrast, containers sourcing water from unprotected dug wells, in Community 2 and 4, had the lowest average bacterial contamination (6.1 log_10_CFU/cm^2^). Prior study found a proportional decline in stored water quality was greater when collected from improved water sources like wells and public standpipes^29^.

### 4.2 Addressing the disconnect between high contamination and household container cleaning practices

Our results suggest a disconnect between biofilm contamination and container cleaning behaviors. Inconsistent container cleaning lessons across programs remain an area for improvement; container cleaning should be a standalone lesson and promoted with consistent messaging across other SWS topics. CHC and CHP training modules differ in origin and structure: CHCs embed container cleaning within broader WASH lessons, while CHPs teach it separately. KIIs and FGDs suggest that an additional focused lesson could improve understanding and adoption by demonstrating cleaning techniques, emphasizing routine cleaning, and highlighting biofilm and recontamination risks. A previous study found that educational interventions encouraged people to keep their containers clean, increased household awareness of container hygiene and reduced biofilm formation^30^. A dedicated container cleaning module with consistent messaging positions it as essential for safe drinking water.

Survey results revealed households often used unreliable indicators, including visible dirt on the container or stored water, to decide when to clean containers. Despite observing the majority of container exteriors being “clean”, exterior container appearance may not necessarily be linked to interior container cleanliness. Observed containers with clean interior and exterior had the lowest average biofilm contamination compared to the other container, however, biofilm contamination was still present. Biofilms are complex microbial communities that adhere to surfaces and can develop rapidly in water storage containers, often without any visual indicators of contamination^10^. Studies have shown that these biofilms can act as reservoirs for pathogenic bacteria, even in containers that appear visibly clean^31^. While keeping containers visibly free of dirt or algae is important, relying on appearance alone could mislead households about true cleanliness.

Cleaning frequency may be a better container cleaning cue, yet defining it remains challenging; AFMAC staff emphasized cleaning before refilling, while program members cited cleaning after unspecified intervals. Other studies assessing factors that impact water quality found that households typically cleaned their transport containers every second or third day^14,32^. These results indicate a need for guidance on cleaning frequencies for both transport and storage containers.

Despite widespread awareness, many respondents reported never formally learning how to clean their containers. Among those who had been taught, most learned from friends or family. AFMAC staff highlighted low levels of membership in CHP and CHC programs due to a lack of incentives to participate. These findings are consistent with CHC programs implemented in Haiti, which found only 4% of non-member households attended CHC meetings, largely due to lack of incentives, negative perception of facilitators, and limited free time^33^. In Haiti, potential incentives to increase CHC participation were the development of social identities and social capital by viewing CHCs as family or “lakou”^33^.

FGD participants shared that limited transportation and demonstration tools hindered outreach, suggesting that bicycles could improve household engagement in remote areas. Evidence from similar interventions supports this approach: in Mozambique, community health workers equipped with bicycles were better able to reach displaced populations and support HIV treatment continuity in hard-to-reach areas^34^. Moreover, a comparative study of CHCs in Rwanda and Zimbabwe emphasized that increased mobility and locally adapted engagement strategies enhanced hygiene behavior change and cost-effectiveness across communities^35^.

Limited access to cleaning supplies due to affordability and prioritization hindered hygienic practices, consistent with systematic review findings citing financial constraints and limited availability as significant barriers to handwashing^36^. In other contexts, CHCs have also demonstrated adaptability in addressing financial constraints to WASH improvements. In Zimbabwe, for example, 28% of CHCs participated in savings and lending initiatives to help households construct latrines, illustrating how financial empowerment can support sanitation efforts^37^. This underscores the need for CHC and CHP programs to improve access to cleaning materials, whether through distribution, supply chains, or subsidies.

Competing domestic household priorities remain a challenge to consistent container cleaning practices. One study found low CHC attendance rates, with only 41% of members attending sessions regularly despite continued program operation^38^. Furthermore, gender norms in study communities indicate that the women head of household bears the responsibility of water collection and container cleaning^40^. As a result, container cleaning may receive less attention while balancing multiple responsibilities, such as childcare and farming. CHC programs in Zimbabwe found that women’s time is a determinant of adopting behavior; specifically, greater agricultural demands allow women heads of households less time to maintain hygiene recommendations^25^. Similarly, in India it was found that education and communication on handwashing only affected women’s behaviors if the intervention only targeted women^39^. Therefore, CHC and CHP programs should target additional household members and addressing factors influencing container cleaning habits, like container type, cleaning difficulty, self-efficacy, commitment, and forgetfulness^15^.

### 4.3 Limitations

This study was limited by its short data collection period, small sample size, and limited number of communities, which may affect generalizability of the findings. Hawthorne bias may have influenced reported cleaning behaviors due to participant awareness of sampling. Future research should involve longer-term studies, larger samples, and strategies to reduce observer bias.

### 4.4 Recommendations

This pilot study highlights the need for additional research on effective, locally acceptable container cleaning methods, emphasizing the value of using available materials and community-specific collaboration, to help evaluate cleaning methods’ long-term effectiveness and further refine household training and workshops. Specifically, further research is needed to examine cleaning frequency across diverse sociocultural and resource-limited settings.

For AFMAC specifically, revising training content to include a dedicated container cleaning module along with embedding it within other SWS lessons, could strengthen its impact. The module should be accessible in multiple languages, incorporate visual training aids, and provide demonstration materials such as cleaning brushes and storage containers. Increasing CHC and CHP member visibility through uniforms and community recognition programs could enhance credibility and encourage greater participation. Additionally, providing transportation for CHC and CHP members could increase program outreach and participation. Lastly, sharing point-of-use water quality responsibilities by redirecting program outreach and sensitizations to additional household members.

## 5.0 CONCLUSION

This mixed-methods study contributes to household water safety by examining container cleaning behaviors and associated challenges within four communities engaged in AFMAC’s CHP and CHC programs. Our findings reveal that while awareness of container cleaning is high among households, consistent and effective practices are limited by material constraints, behavioral norms, and variability in training content across CHC and CHP platforms. From our findings, high bacterial contamination were observed in storage containers, underscoring the gap between knowledge and practice, and highlighting container cleaning as a critical pathway for post-source water recontamination. Our findings support progress towards SGD 6 through the promotion of safely managed drinking water and reducing the risk of waterborne illnesses at the household level.

The study also emphasizes the importance of contextually grounded, community-responsive approaches in WASH programming. AFMAC’s program design, rooted in community participation and peer engagement, demonstrates promising strategies for behavior change. However, the lack of a standardized, standalone module on container cleaning across all training formats, the prevalent reliance on visual cues as proxies for microbial safety, and the disproportionate responsibility placed on women to manage household water hygiene collectively underscore critical gaps in current programming that warrant targeted intervention.

To address these gaps, future interventions should prioritize the design and implementation of standardized modules on container cleaning, ensure reliable access to essential cleaning materials, and broaden the scope of behavior change efforts to actively engage all household members in the community. Further research is needed to evaluate the long-term effectiveness of these interventions and to examine cleaning frequency across diverse sociocultural and resource-limited settings.

Advancing container hygiene through integrated, adaptive, and scalable approaches is essential for reducing public health risks from unsafe water storage and improving point-of-use water quality.

## DECLARATION

### Ethics approval and consent to participate

The study protocol was approved by the Lehigh University Institutional Review Board (Protocol# 2137976-1) and the ERES Converge Institutional Review Board in Zambia (Reference# 2024-Feb- 015), after both PI and local Co-PI were registered as researchers with the National Health Research Commission in Zambia. Clinical trial number: not applicable.

Staff familiar with the communities and local language were hired as research assistants (RA). Over five days, RAs were trained by Lehigh staff on ethics, informed consent, FGD questions and moderation, household survey tools, water quality testing, and biofilm sample collection. Verbal consent was obtained from each household before administering the survey to an adult over age 18 (preferably female head of household). Verbal or written consent was obtained from each FGD participant and KII participant, respectively.

## Consent for publication

Not applicable

## Availability of data and materials

The datasets for the household survey and biofilm contamination can be found in an online repository at: https://osf.io/dtp7k/?view_only=5fa57ed63c17467d8dfe40f26be757a0

## Authors’ contributions

Tracy Zhang: Conceptualization, Methodology, Investigation, Formal Analysis, Validation, Data curation, Writing - Original Draft, Writing - Review & Editing, Visualization

Yanwen Li: Validation, Formal Analysis, Data curation, Writing - Original Draft, Writing - Review & Editing

Nathan Nonde: Investigation, Writing - Original Draft

Elijah Mutafya: Project administration, Writing - Original Draft, Writing - Review & Editing Gabrielle String: Conceptualization, Resources, Validation, Writing - Original Draft, Writing - Review & Editing, Supervision, Funding acquisition

## Supporting information

Supplemental Background S1

## Data Availability

All data produced are available online at

https://osf.io/dtp7k/?view_only=5fa57ed63c17467d8dfe40f26be757a0

## Acknowledgments

The authors would like to thank CAWST for funding and coordination, especially Marcio Botto, Toby Gould, and Camille Zimmer. We appreciate the work of our local research assistants, Samson Sakala and Esau Banda. Lastly, we are grateful to all of our study participants in Ndola for giving us their time.

## Authors’ Information

Tracy Zhang

College of Health, Lehigh University Bethlehem, PA, USA

Yanwen Li

College of Health, Lehigh University Bethlehem, PA, USA

Nathan Nonde

Africa MANZI Centre 2J9V+JQX, Ndola, Zambia

Elijah Mutafya

Africa MANZI Centre 2J9V+JQX, Ndola, Zambia

*Gabrielle String

Department of Civil and Environmental Engineering, P.C. Rossin College of Engineering and Applied Sciences, Lehigh University

Department of Population Health, College of Health, Lehigh University Bethlehem, PA, USA

## REFERENCES

(1) WHO. State of the World’s Drinking Water: An Urgent Call to Action to Accelerate Progress on Ensuring Safe Drinking Water for All; World Health Organization, 2022. https://www.who.int/publications-detail-redirect/9789240060807 (accessed 2023-09-24).

(2) Cassivi, A.; Tilley, E.; Waygood, E. O. D.; Dorea, C. Household Practices in Accessing Drinking Water and Post Collection Contamination: A Seasonal Cohort Study in Malawi. Water Research 2021, 189, 116607. 10.1016/j.watres.2020.116607.

(3) Rufener, S.; Mäusezahl, D.; Mosler, H.-J.; Weingartner, R. Quality of Drinking-Water at Source and Point-of-Consumption - Drinking Cup As a High Potential Recontamination Risk: A Field Study in Bolivia. *Journal of Health*, Population and Nutrition 2010, 28 (1), 34–41. 10.3329/jhpn.v28i1.4521.

(4) World Health Organization Regional Office for the Western. Household Water Treatment and Safe StoragelJ: Manual for the Participant; WHO Regional Office for the Western Pacific, 2013.

(5) CDC. About Global Water, Sanitation and Hygiene (WASH). Global Water, Sanitation, and Hygiene (WASH). https://www.cdc.gov/global-water-sanitation-hygiene/about/index.html (accessed 2024-06-30).

(6) *Goal 6 | Department of Economic and Social Affairs*. https://sdgs.un.org/goals/goal6 (accessed 2025-06-12).

(7) *Goal 3 | Department of Economic and Social Affairs*. https://sdgs.un.org/goals/goal3#targets_and_indicators (accessed 2025-06-12).

(8) String, G.; Domini, M.; Mirindi, P.; Brodsky, H.; Kamal, Y.; Tatro, T.; Johnston, M.; Badr, H.; Lantagne, D. Efficacy of Locally-Available Cleaning Methods in Removing Biofilms from Taps and Surfaces of Household Water Storage Containers. *npj Clean Water* 2020, 3 (1), 1–11. 10.1038/s41545-020-0061-y.

(9) Petrova, O. E.; Sauer, K. Escaping the Biofilm in More than One Way: Desorption, Detachment or Dispersion. Current Opinion in Microbiology 2016, 30, 67–78. 10.1016/j.mib.2016.01.004.

(10) Jagals, P.; Jagals, C.; Bokako, T. C. The Effect of Container-Biofilm on the Microbiological Quality of Water Used from Plastic Household Containers. Journal of Water and Health 2003, 1 (3), 101–108. 10.2166/wh.2003.0012.

(11) Meierhofer, R.; Wietlisbach, B.; Matiko, C. Influence of Container Cleanliness, Container Disinfection with Chlorine, and Container Handling on Recontamination of Water Collected from a Water Kiosk in a Kenyan Slum. Journal of Water and Health 2019, 17 (2), 308–317. 10.2166/wh.2019.282.

(12) String, G. M.; Domini, M.; Badr, H.; Brodsky, H.; Kamal, Y.; Tatro, T.; Johnston, M.; Ogudipe, A.; Nha Vu, T.; K. Wolfe, M.; S. Lantagne, D. Efficacy of Locally-Available Cleaning Methods and Household Chlorination at Inhibiting Biofilm Development in Jerricans Used to Store Household Drinking Water. Environmental Science: Water Research & Technology 2021, 7 (2), 367–383. 10.1039/D0EW00748J.

(13) Judah, L. A.; Andriambololonirina, C.; Rakotoarisoa, L.; Barrett, L. J. P.; Khaliq, M.; Mihelcic, J. R.; Cunningham, J. A. Occurrence and Mitigation of Bacterial Regrowth in Stored Household Water in Eastern Coastal Madagascar. Water 2024, 16 (11), 1592. 10.3390/w16111592.

(14) Gärtner, N.; Germann, L.; Wanyama, K.; Ouma, H.; Meierhofer, R. Keeping Water from Kiosks Clean: Strategies for Reducing Recontamination during Transport and Storage in Eastern Uganda. Water Research X 2021, 10, 100079. 10.1016/j.wroa.2020.100079.

(15) Stocker, A.; Mosler, H.-J. Contextual and Sociopsychological Factors in Predicting Habitual Cleaning of Water Storage Containers in Rural Benin. Water Resources Research 2015, 51 (4), 2000–2008. 10.1002/2014WR016005.

(16) Bae, S.; Lyons, C.; Onstad, N. A Culture-Dependent and Metagenomic Approach of Household Drinking Water from the Source to Point of Use in a Developing Country. Water Research X 2019, 2, 100026. 10.1016/j.wroa.2019.100026.

(17) Budeli, P.; Moropeng, R. C.; Mpenyana-Monyatsi, L.; Momba, M. N. B. Inhibition of Biofilm Formation on the Surface of Water Storage Containers Using Biosand Zeolite Silver-Impregnated Clay Granular and Silver Impregnated Porous Pot Filtration Systems. PLoS One 2018, 13 (4), e0194715. 10.1371/journal.pone.0194715.

(18) Kanyunge, C. M. Integrating Household Water Treatment and Safe Storage Practices in Zambia’s National Water Policy for Effective Regulation, Evaluation and Sustainable Water Provision. Master Thesis, PAUWES, 2020. http://repository.pauwes-cop.net/handle/1/410 (accessed 2024-06-30).

(19) UNICEF WHO Joint Monitoring Program. The WHO/UNICEF Joint Monitoring Programme (JMP) Data, 2022. https://washdata.org/data/household#!/ (accessed 2024-06-30).

(20) Peletz, R.; Simunyama, M.; Sarenje, K.; Baisley, K.; Filteau, S.; Kelly, P.; Clasen, T. Assessing Water Filtration and Safe Storage in Households with Young Children of HIV-Positive Mothers: A Randomized, Controlled Trial in Zambia. PLOS ONE 2012, 7 (10), e46548. 10.1371/journal.pone.0046548.

(21) Peletz, R.; Simuyandi, M.; Simunyama, M.; Sarenje, K.; Kelly, P.; Clasen, T. Follow-Up Study to Assess the Use and Performance of Household Filters in Zambia. Am J Trop Med Hyg 2013, 89 (6), 1190–1194. 10.4269/ajtmh.13-0054.

(22) CAWST. *Safe Drinking Water Storage and Handling | WASH Resources*. WASH Resources. https://washresources.cawst.org/en/module/ce8104f2/safe-drinking-water-storage-and-handling (accessed 2025-03-05).

(23) *AFMAC - Community WASH Promotion*. CAWST | Training. https://www.cawst.training/event/afmac_-_community_wash_promotion102248322 (accessed 2025-06-12).

(24) CAWST. *Biofilm Infographic: Are you protecting those you serve? | WASH Resources*. WASH Resources. https://washresources.cawst.org/en/resources/f378c5cc/biofilm-infographic (accessed 2025-03-05).

(25) Waterkeyn, J.; Cairncross, S. Creating Demand for Sanitation and Hygiene through Community Health Clubs: A Cost-Effective Intervention in Two Districts in Zimbabwe. Social Science & Medicine 2005, 61 (9), 1958–1970. 10.1016/j.socscimed.2005.04.012.

(26) *Heterotrophic Plate Counts and Drinking-Water Safety: The Significance of HPCs for Water Quality and Human Health*; Bartram, J., Cotruvo, J. A., Exner, M., Fricker, C., Glasmacher, A., Eds.; Emerging issues in water and infectious disease series; IWA Publ: London, 2003.

(27) Tolley, E. E. *Qualitative Methods in Public Health: A Field Guide for Applied Research*; John Wiley & Sons, Incorporated: Newark, UNITED STATES, 2016.

(28) Carabin, A.; Cassivi, A.; Dorea, C.; Rodriguez, M.; Huot, C. Heterotrophic Plate Counts (HPC) in Drinking Water Distribution Systems: A Comprehensive Review and Meta-Analysis. Water Quality Research Journal 2024, wqrj2024027. 10.2166/wqrj.2024.027.

(29) Wright, J.; Gundry, S.; Conroy, R. Household Drinking Water in Developing Countries: A Systematic Review of Microbiological Contamination between Source and Point-of-Use. Tropical Medicine & International Health 2004, 9 (1), 106–117. 10.1046/j.1365-3156.2003.01160.x.

(30) Nala, N. P.; Jagals, P.; Joubert, G. The Effect of a Water-Hygiene Educational Programme on the Microbiological Quality of Container-Stored Water in Households. Water SA 2003, 29 (2), 171–176. 10.4314/wsa.v29i2.4852.

(31) Makokove, R.; Macherera, M.; Kativhu, T.; Gudo, D. F. The Effect of Household Practices on the Deterioration of Microbial Quality of Drinking Water between Source and Point of Use in Murewa District, Zimbabwe. Journal of Water and Health 2022, 20 (3), 518–530. 10.2166/wh.2022.251.

(32) Meierhofer, R.; Rubli, P.; Dreyer, K.; Ouma, H.; Wanyama, K.; Peter-Varbanets, M. Membrane Filtration Reduces Recontamination Risk in Chlorinated Household Water Containers. 2017.

(33) Brooks, J.; Adams, A.; Bendjemil, S.; Rosenfeld, J. Putting Heads and Hands Together to Change Knowledge and Behaviours: Community Health Clubs in Port-Au-Prince, Haiti. Waterlines 2015, 34 (4), 379–396.

(34) *-Decreasing the Distance to Health: ICAP Partners with Bicycle Company in Mozambique to Mobilize HIV Services*. https://icap.columbia.edu/news-events/decreasing-the-distance-to-health-icap-partners-with-bicycle-company-in-mozambique-to-mobilize-hiv-services/ (accessed 2025-04-08).

(35) Waterkeyn, J.; Matimati, R.; Muringaniza, A.; Chigono, A.; Ntakarutimana, A.; Katabarwa, J.; Bigirimana, Z.; Pantoglou, J.; Waterkeyn, A.; Cairncross, C.; Cairncross, A. Comparative Assessment of Hygiene Behaviour Change and Cost-Effectiveness of Community Health Clubs in Rwanda and Zimbabwe.; Rozman, U., Ed.; IntechOpen, 2019.

(36) Ezezika, O.; Heng, J.; Fatima, K.; Mohamed, A.; Barrett, K. What Are the Barriers and Facilitators to Community Handwashing with Water and Soap? A Systematic Review. PLOS Glob Public Health 2023, 3 (4), e0001720. 10.1371/journal.pgph.0001720.

(37) Murakwani, P. N.; Sibanda, W.; Dube, S. B.; Weber, N. Community Health Clubs Improve Latrine Construction through Savings, Lending, and Income-Generating Activities. *Journal of Water*, Sanitation and Hygiene for Development 2022, 12 (2), 227–236. 10.2166/washdev.2022.084.

(38) Waterkeyn, J.; Waterkeyn, A.; Uwingabire, F.; Pantoglou, J.; Ntakarutimana, A.; Mbirira, M.; Katabarwa, J.; Bigirimana, Z.; Cairncross, S.; Carter, R. The Value of Monitoring Data in a Process Evaluation of Hygiene Behaviour Change in Community Health Clubs to Explain Findings from a Cluster-Randomised Controlled Trial in Rwanda. BMC Public Health 2020, 20 (1), 98. 10.1186/s12889-019-7991-7.

(39) Cairncross, S.; Shordt, K.; Zacharia, S.; Govindan, B. K. What Causes Sustainable Changes in Hygiene Behaviour? A Cross-Sectional Study from Kerala, India. Soc Sci Med 2005, 61 (10), 2212–2220. 10.1016/j.socscimed.2005.04.019.

(40) Leontsini, E.; Maloney, S.; Ramírez, M.; Rodriguez, E.; Gurman, T.; Ballard Sara, A.; Hunter, G. C. A Qualitative Study of Community Perspectives Surrounding Cleaning Practices in the Context of Zika Prevention in El Salvador: Implications for Community-Based Aedes Aegypti Control. BMC Public Health 2020, 20 (1), 1385. 10.1186/s12889-020-09370-5.

