## Supplemental Background S1 for "Integrating container cleaning practices into a Zambian Community Health Club program - evidence from a pilot study"

### FIGURES

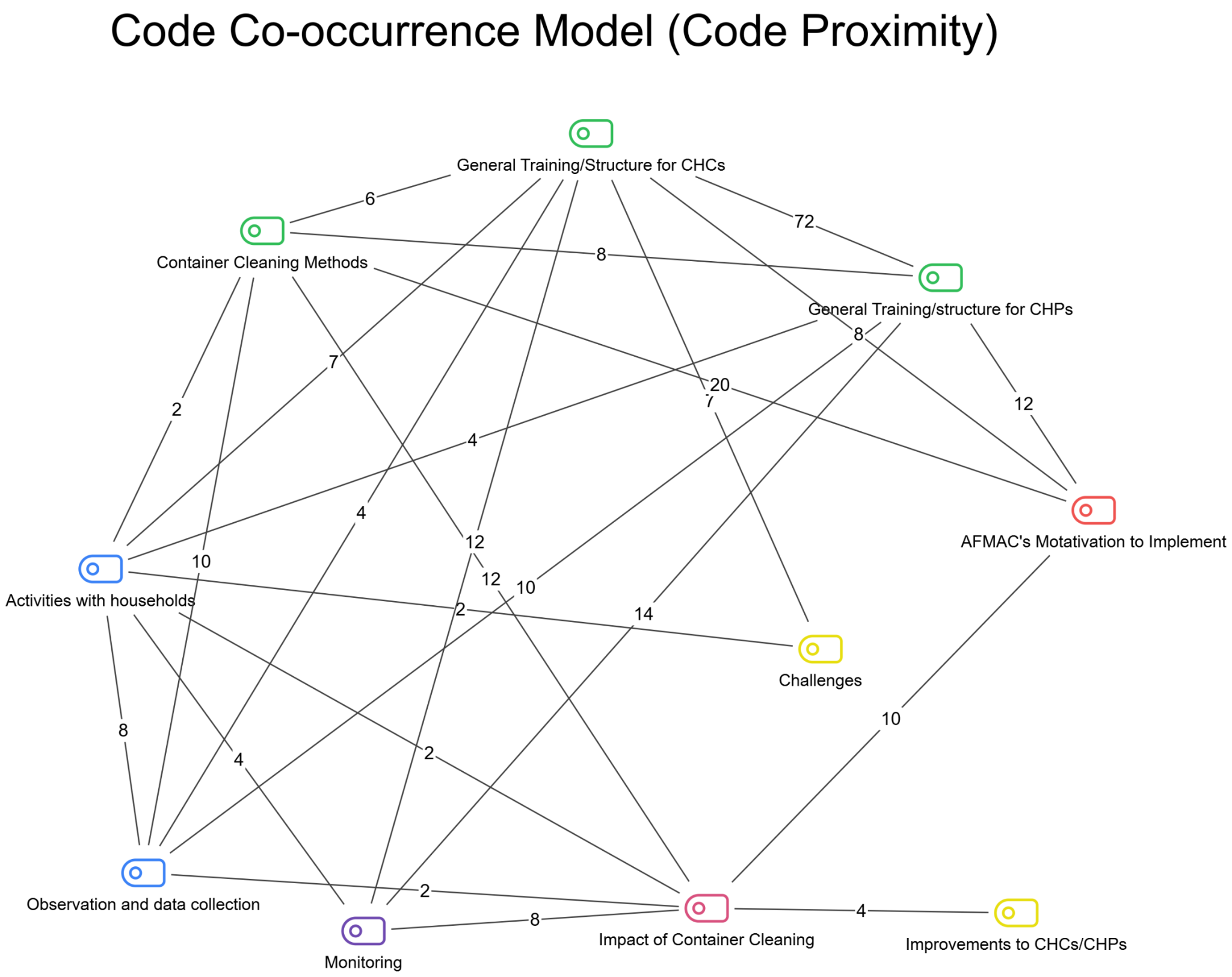

**Figure S1.** KII Code Co-occurrence Model - depicts the proximity of coded interview segments to each other representing how often informants discussed topics in relation to each other. Model created in MAXQDA for 3 minimum co-occurrences, a maximum distance of 1 paragraph apart. Line labels represent the frequency of co-occurrences between codes. Note, there are multiple sub-codes depicted for the categories of *AFMAC training* (green) , *CHC/CHP engagement with communities* (blue), and *Program feedback* (yellow).

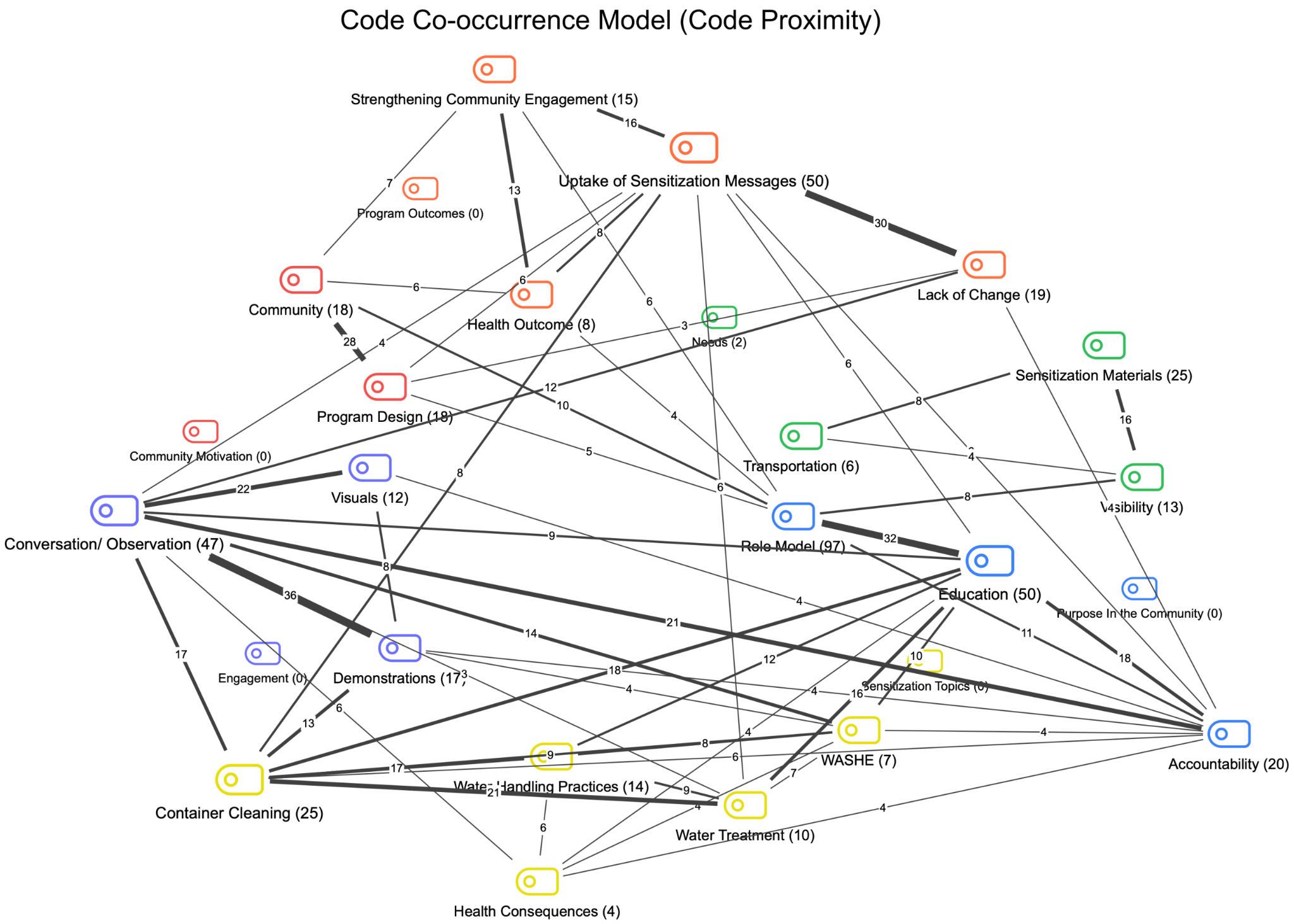

**Figure S2.** FGD Code Co-occurrence Model - depicts the proximity of coded transcript segments to each other representing how often participants discussed topics in relation to each other. Model created in MAXQDA for 3 minimum co-occurrences, a maximum distance of 1 paragraph apart. Line labels represent the frequency of co-occurrences between codes. Note, there are multiple sub-codes depicted for the categories of *CHC/CHP needs* (green) , *Purpose in the community* (blue), *Community motivations* (red), *Engagement tools* (purple), *Program outcomes* (orange), and *Sensitization topics* (yellow).

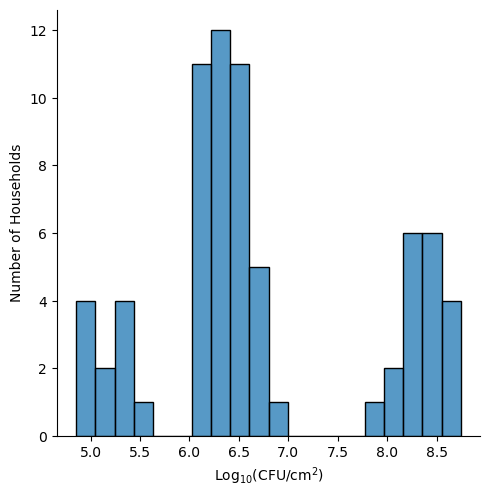

**Figure S3. Log Transformed Distribution of Bacterial Contamination of All Households** - Barplot depicting the frequency of households at each bacterial contamination level. There is a bimodal distribution with two distinct groups of contamination.

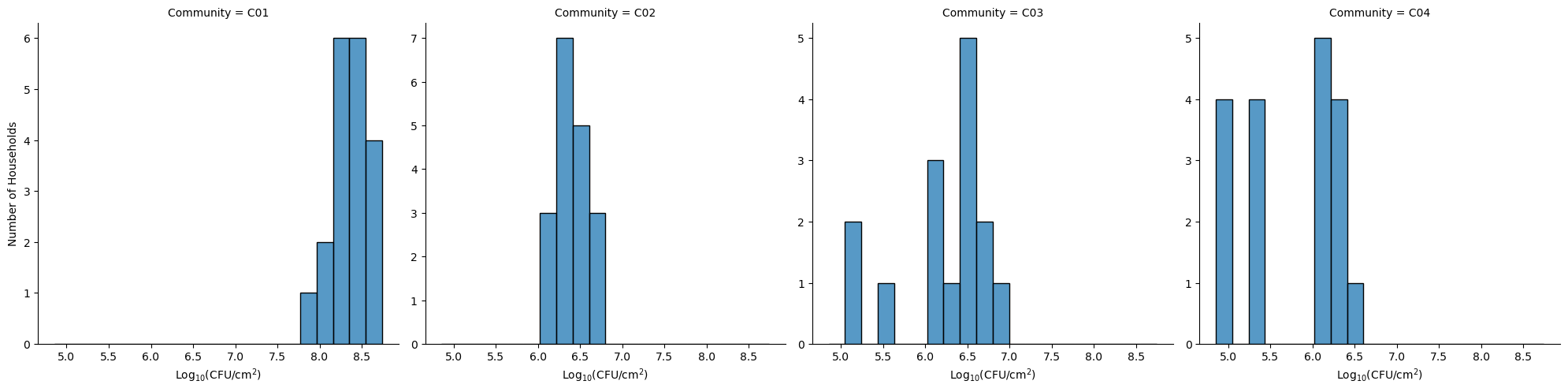

**Figure S4. Log Transformed Distribution of Bacterial Contamination By Community** - Barplot depicting the frequency of households by contamination level within each community. Community 1 shows a left skewed distribution. Community 2 shows symmetrical distribution with contamination centered around 6-7 CFU/cm^2^. Community 3 and 4 have more variable distributions with a greater range in contamination levels.

### TABLES

**Table S1. Additional Household Demographic and Water Collection and Treatment.** Data from household surveys (n=73) is presented alongside the mean biofilm log10 HPC CFU/cm^2^ for the corresponding household survey category.

| Variables | | Households (n=73) | | Container biofilm^a^  Mean (range) | |
| --- | --- | --- | --- | --- | --- |
| N(%) Respondent's Sex | | | | | |
|  | Male respondents | 18 | (25) | 7.8 | (5.5-8.5) |
|  | Female respondents | 55 | (75) | 6.5 | (4.9-8.7) |
| N(%) Respondent's age range | | | | | |
|  | 18-24 | 9 | (12) | 6.4 | (4.9-8.3) |
|  | 25-49 | 45 | (62) | 6.8 | (4.9-8.7) |
|  | 50-64 | 16 | (22) | 6.8 | (5.0-8.7) |
|  | 65+ | 3 | (4.1) | 6.7 | (5.5-8.3) |
| N(%) Respondent's highest level of education completed, n=64 | | | | | |
|  | Primary | 24 | (38) | 6.3 | (4.9-8.5) |
|  | Secondary | 40 | (63) | 7.0 | (4.9-8.7) |
| N(%) Respondents with Head of Household (HOH) that can read and write | | | | | |
|  | Female HOH that can read and write, n=72 | 57 | (79) | 6.8 | (4.9-8.7) |
|  | Male HOH that can read and write, n=65 | 61 | (94) | 6.8 | (4.9-8.7) |
| Mean (range) number of people per household | | 5.4 | (1-11) | - | - |
| N(%) Someone in home has had diarrhea in last week | | 28 | (38) | 6.4 | (5.0-8.4) |
|  | Mean (range) number of people with diarrhea in the last week, n=28 | 1.5 | (1-3) | - | - |
| Median (range) minutes spent collecting water per trip, n=72 | | 10 | (2-30) | - | - |
| N (%) Sand filter for treating water, n=7 | | 6 | (86) | 6.6 | (6.4-6.8) |

*a Three households had plate growth that exceeded the maximum detection limit and were excluded*

*from mean HPC concentrations (n=70)*

** p-value <0.05; ** p-value <0.01; *** p-value <0.001*

**Table S2. Water Quality Parameters**. Data from household surveys (n=73) is summarized for the corresponding household survey category. The mean or median values for water quality parameters, along with their corresponding ranges are presented.

| Variables | Households (n=73) | |
| --- | --- | --- |
| mean (range) drinking water sample pH | 6.6 | (5.3- 8.6) |
| mean (range) drinking water sample temperature,°C | 25.1 | (13- 52) |
| median (range) drinking water sample total dissolved solid (TDS), ppm | 60 | (12- 317) |
| median (range) drinking water sample electrical conductivity (EC), uS/cm | 78 | (17- 445) |

### BACKGROUND

#### Background S1: Cleaning Strategies and Materials

Various industries have implemented pipe cleaning and disinfection strategies to control biofilm growth in contact with potable water ^1,2^. For large household storage tanks, utilized for prolonged storage of water, common container cleaning methods include using automatic equipment (e.g., mechanical scrubbers and vacuum cleaners) or manual tools (e.g., mops and brooms) ^3^. Many studies have found frequent tank cleanings, prior to water refill ^3^ or at least three times a year ^4^, can improve bacterial contamination ^5^.

The materials used to clean and disinfect household water storage containers differ for container type and vary in effectiveness at reducing recontamination. The use of chlorine-based disinfectants has been shown in studies to be effective at reducing bacterial contamination in water storage containers ^6–10^. However, chlorine-based disinfectants are not locally available and affordable to communities in low- and middle-income countries (LMICs). Alternatively, sand is a locally available and inexpensive material used to clean jerrycans. A field study in Uganda found the use of sand decreased recontamination with E. coli but did not reduce the total coliform levels. Sand can scratch the inner walls of containers and can increase the chlorine demand ^11^. For improved containers with a wide opening and a spigot, the use of brushes to clean resulted in lower counts of total coliform and reduced E. coli recontamination ^11^. Similarly, laboratory-based research on jerrican cleaning found *E. coli* biofilm growth was inhibited using sodium hypochlorite alone and in combination with rocks, although the use of rocks may also promote biofilm growth by scratching container surfaces ^9^. Overall, the field evidence on household water storage container cleaning is limited, and laboratory-tested methods do not guarantee to be best practices in varied real-world settings.

#### Background S2: Cleaning Frequency Studies

Meierhofer *et al.* found that containers disinfected with chlorine along with maintaining high levels of Free Residual Chlorine (FRC) and frequent container cleaning were associated with reduced recontamination during 24 hour storage^8^. While it was reported households cleaned their containers once a week, it is unclear how much more frequent cleaning must occur to be effective ^8^. In a cross-sectional study, it was found that a majority of households cleaned their containers before they refilled them and expected to refill their containers within three days ^12^. While there is no widely accepted definition of how frequently containers need to be cleaned to prevent biofilm formation, it was found that water storage for more than 3 days without cleaning increased health risks ^13^. Budeli *et al.* found that biofilm formation on the surface of plastic storage containers can occur within 24 hours with untreated water ^14^. Specifically, they found biofilm formation occurred 14-21 days after water treatment by the silver-impregnated porous pot system and 3-14 days after water treatment by the bio-sand zeolite silver-impregnated clay granular filter. They suggested disinfection should occur within 14 and 7 days, respectively, to prevent buildup of biofilms ^14^.

#### Background S3: HWTS in Zambia

A randomized control trial of Zambian households with children under 2 years found household use of a water filter and safe storage was highly effective in improving water quality and preventing diarrhea; households in the intervention arm had significantly better microbiological water quality than those who did not (p<0.001) ^15^. After one year, filters continued to provide high-quality water, removing 99% of fecal indicator bacteria, and 75% of households had stored water of better quality than the source ^16^.

#### Background S4: CHC/CHP Program Structure

CHPs are volunteers individually trained by AFMAC staff using CAWST’s guidance ^17,18^. Training focuses on practical, household-level interventions CHPs can use to promote WASH-related topics within their community. CHPs are requested to visit a few households per week to promote WASH topics and identify any challenges to be reported back to AFMAC. There are about 1,200 CHPs within the communities.

CHCs are a group of self-sustaining community members dedicated to conducting WASH-related training and discussing challenges faced within the community. Community members take up leadership roles consisting of chairperson, vice chairperson, secretary, treasurer, and facilitator. Members undergo a six-month interactive training program covering 26 lessons on hygiene, sanitation, and water treatment using material adapted from Africa Ahead, a non-profit organization in Zimbabwe ^19^. Each club selects a facilitator from within the community to be trained by AFMAC and lead CHC sessions. CHCs have been proven to be scalable and cost-effective, as seen in Haiti, where CHC programs expanded from six to 66 communities between 2012 and 2015 while maintaining knowledge retention and reducing open defecation^20^.

#### Background S5: Top reported health concerns by community

Kaloko is a periurban community with a population of 18,454 people in 3,075 households. Mwange is a rural community of 1,800 people in 310 households. George Compound is a periurban community of 10,186 people and 1,600 households. Chilibi Village is a rural community of 88 households.

Kaloko ^21^

1. Malaria
2. Diarrhea
3. Respiratory tract infections

Mwange ^22^

1. Malaria
2. Diarrhea
3. Respiratory tract infections

Chibili ^23^

1. Malaria
2. Diarrhea
3. Respiratory tract infections
4. STIs

George ^24^

1. Malaria
2. Diarrhea
3. Respiratory tract infections
4. STIs

### RESULTS

#### Results S1: Roles of AFMAC vs CHCs & CHPs

AFMAC leadership is responsible for designing, managing, and overseeing the CHC and CHP programs, making sure that training materials, monitoring tools, and intervention strategies align with public health goals. Their role is mainly administrative and strategic, focusing on setting program guidelines, distributing resources, and addressing logistical challenges like funding and evaluation.

In contrast, CHC and CHP leaders work directly within communities, engaging with households to put these interventions into practice. CHC leaders guide group-based learning, encouraging peer accountability and collective behavior change, while CHP leaders take a more hands-on approach, visiting homes and providing personalized guidance on container cleaning and water safety. While AFMAC leadership sets the overall structure and strategic direction, CHC and CHP leaders bring these policies to life at the community level. This distinction highlights the need for strong support at both levels to ensure long-term improvements in household water storage practices.

#### Results S2: Behavior Change Through Demonstrations and Continuous Engagement

Demonstrations were repeatedly identified as a cornerstone of the sensitization process. Facilitators often cleaned visibly dirty containers during household visits to illustrate proper techniques. One participant shared, “I noticed the container had a lot of algae, so I asked for a sponge, some sand, soap, and luckily chlorine, which I used to clean it. I told her this is how a storage container should be cleaned” (FGD C02). Demonstrations allow households to “...see the whole process, they understand why it is important”. Practical container training resonates strongly with community members, who often learn best through visual and experiential methods. Demonstrations are often paired with conversations, emphasizing the need for proper cleaning frequency. For example, a facilitator explained, “We advise them to clean the container before every refill and how to take care of the filter” (FGD C01). However, continuous engagement is necessary to sustain behavior change. Facilitators have also noted that some households reverted to previous habits due to competing priorities or a lack of resources, which highlights the need for follow-ups and reinforcement (FGD C03).

#### Results S3: Stored Drinking Water Quality

Physical chemical water quality samples were collected from 73 households. Average drinking water pH was 6.57 and the average temperature was 25.1 ºC (Table S2). The median total dissolved solids (TDS) was 59.6 ppm, ranging from 11.5-317 ppm (Table S2). The median electrical conductivity (EC) was 78.2 μS/cm and ranged from 17.4-445 μS/cm (Table S2).

### DISCUSSON

#### Discussion S1: Strategies to Promote Safe Water Storage and Cleaning in Community Health Programs

AFMAC’s decision to incorporate container cleaning into its programming was influenced by recurring health concerns reported from the communities it served. All respondents recognized the need and importance of container cleaning. Additionally, the top reported health risks of unclean containers were diarrhea, cholera, and stomachaches. AFMAC staff echoed these findings, noting higher diarrhea incidences during the rainy season, highlighting the need for improved hygiene and water storage practice interventions. Furthermore, AFMAC noticed that family traditions and local customs such as only rinsing containers or cleaning when visibly dirty, influenced household water quality. Such beliefs are common; for instance, in southern Sri Lanka, only one of eleven perceived causes of diarrhea was attributed to dirty water, highlighting a general lack of awareness about the link between water quality and health ^25^. AFMAC’s container cleaning initiatives were shaped by both empirical observations of public health risks and the articulated needs of the communities, reinforcing the importance of responsive and participatory program design.

Training plays a critical role in equipping program members with the necessary skills to lead sensitization efforts effectively. AFMAC employs two different training strategies: CHC members participate in a multi-session curriculum covering a range of WASH-related topics, while CHP training is more hands-on, focusing on household-level interventions. Both programs utilize community engagement to foster strong relationships and active participation. Critically, AFMAC creates community ownership of these programs by entrusting local leaders to enforce both group-based CHC activities and individualized CHP visits to ensure that households receive targeted support. CHCs foster a sense of accountability among members, encouraging peer monitoring and leading by example, while CHPs provide tailored guidance during home visits. Evidence from Rwanda supports the long-term effectiveness of CHCs in sustaining improved WASH behaviors, with participants maintaining better sanitation and hygiene practices even three years after program implementation ^26^. Additionally, in Zimbabwe, large-scale behavior change was observed as over 24,000 people participated in CHC-led hygiene sessions at a low cost per participant, reinforcing the approach as both impactful and cost-effective ^19^.

During container cleaning sensitizations, engagement tools such as demonstrations and visual aids ensure community participation across literacy levels. This technique has been implemented in other studies where demonstrations and visual aids have improved handwashing knowledge and behavior in school settings ^27,28^. In Zimbabwe CHCs, the use of visual aids and participatory activities did not require certain levels of literacy to be successful ^19^. Interviewees emphasized that tailoring training materials to reflect local customs, such as traditional cleaning with ash or sand, has enhanced program relevance and improved adoption. During demonstrations, members guided households step-by-step in cleaning containers while adapting to materials readily available at home, allowing households to observe and replicate. Many facilitators relied on visual aids like lesson cards paired with conversation to make the information more accessible. Program members reported that these interactive approaches improved their understanding and encouraged greater adoption of best practices. The success of CHC training in sustaining engagement has been documented in Burkina Faso, where 650 CHC members successfully completed a 22-week WASH course, demonstrating retention and long-term participation ^29^.

In our study, KII and FGD participants discussed scrubbing containers with soap, sand, or ash and disinfecting with chlorine afterwards when available. However, surveyed households predominantly used water and a sponge/brush and may have used soap. This is different from a similar study where households cleaned jerrycans with sand (82%), soap (48%) and sponges (57%) ^30^. These cleaning methods have shown to reduce biofilms and be protective of water quality in previous studies. For instance, a laboratory study assessing cleaning methods at inhibiting biofilm development in jerricans found biofilm was inhibited when jerrycans were cleaned with chlorine and rocks ^9^. One field study found factors with a protective effect on water quality were chlorination, cleaning containers with sand, and access to soap for handwashing ^30^; another found that cleaned Maji Safi containers that had been disinfected with chlorine had significantly lower levels of recontamination with *E.coli* and total coliform compared to unclean jerricans ^8^. These studies suggest chlorine as an additional step can be protective for water quality and should be emphasized by container cleaning lessons with CHC and CHP members.

Implementing monitoring and feedback strategies informed AFMAC of the overall status of programs and community members of individual and peer progress. These strategies allow AFMAC to monitor disease prevalence while also collecting feedback on additional WASH topics to support community needs. AFMAC staff and program members have expressed positive and negative behavioral and health outcomes from the program. Many reported noticeable reductions in diarrhea and other waterborne diseases as well as adoption of recommended container cleaning practices. Similar findings have been observed in Zambia, where individuals who completed CHC training were over five times more likely to drink safe water and more than seven times more likely to store drinking water safely compared to those who were not trained ^31^. However, focus group discussions have highlighted a challenge with households refusing to adopt behavior and maintain container cleaning practices.

### MATERIALS AND METHODS

### Methods S1: Data Collection Tools

#### Methods S1.1: Random Sampling Approach

Random route sampling is a sampling method where enumerators complete a predetermined walking path and approach every Nth household to conduct a survey. The walk continues for as long as the enumerator completes the target number of N interviews ^32^.

#### Methods S1.2: Key Informant Interview Guide

—----- Overview —-----

1. To start, can you tell me about the work you do related to community health clubs and community health promoters?
   a. Is there a part in your work that involves container cleaning or container cleaning sensitizations? Explain. (*e.g., Promoting container cleaning practices in training of CHCs or CHPs or in sensitization….*)
2. Why were the communities that have CHC/CHP programs chosen?
3. What are some reasons why CHPs and CHCs were implemented? (*In general*)
4. How frequently do training for CHCs and CHPs take place, and what factors influence their scheduling?
5. What does the training process for CHPs and CHCs entail?
6. Describe the current resources available for CHCs and CHPs (*e.g., staff, material, time*). If they differ, how do they differ between communities?
7. What were some challenges, if any, with conducting the training for CHCs and CHPs?
8. Are CHCs and CHPs promoting container cleaning practices in their sensitization?
   a. If so, please describe how they are promoting container cleaning practices in their sensitizations.

—----- Community and Behavior —-----

9. Have you noticed differences in the way communities clean containers? (*differ by community?*)
   a. Follow-up if yes →What differences are those?
10. In the communities you have worked with, what were container cleaning practices/behavior before the implementation of CHCs or CHPs?
    a. Follow-up with "Did you notice changes in practice after the implementation of CHPs and CHCs? What were these changes?
11. How do you ensure communities continue to engage with the CHC and CHP?

—----- Opinions —-----

12. How do you believe the container cleaning sensitizations have impacted the community?
    a. What are some ways that CHPS and CHCs can be improved?
13. Is there anything else about CHPs and CHCs that you would like to share that we have not already discussed?
14. Are there any ideas/thoughts about container cleaning that you would like to share that we have not already discussed?

#### Methods S1.3: Focus Group Discussion Guide

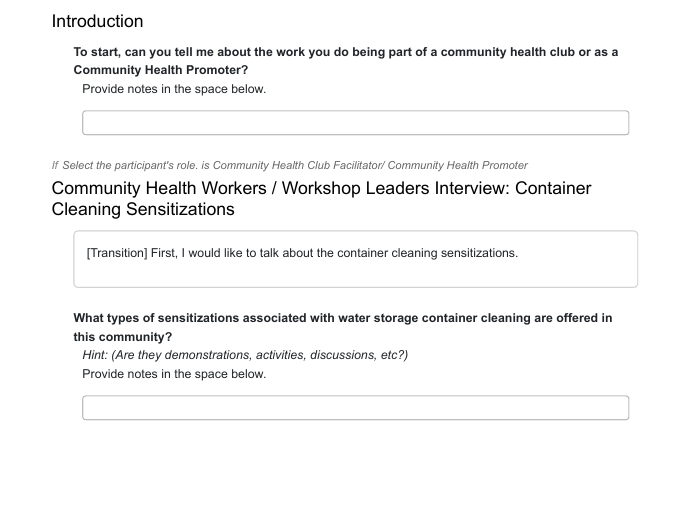

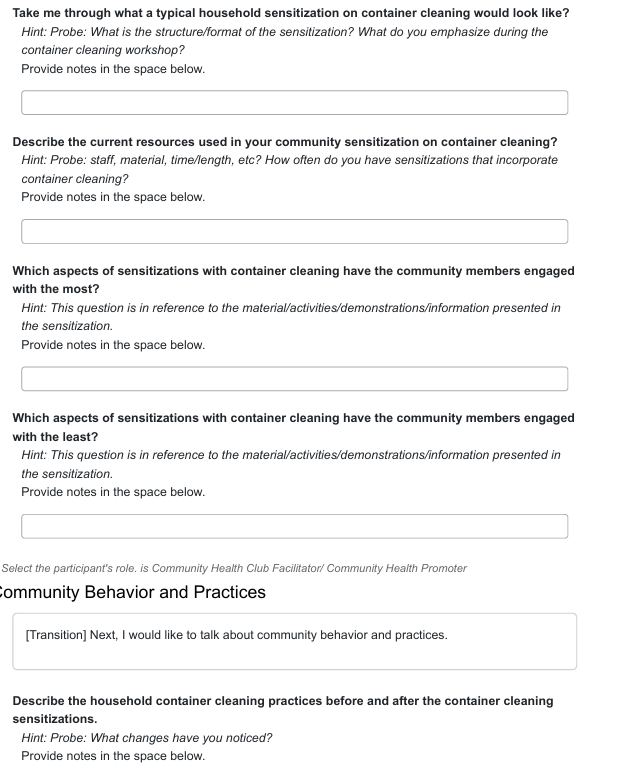

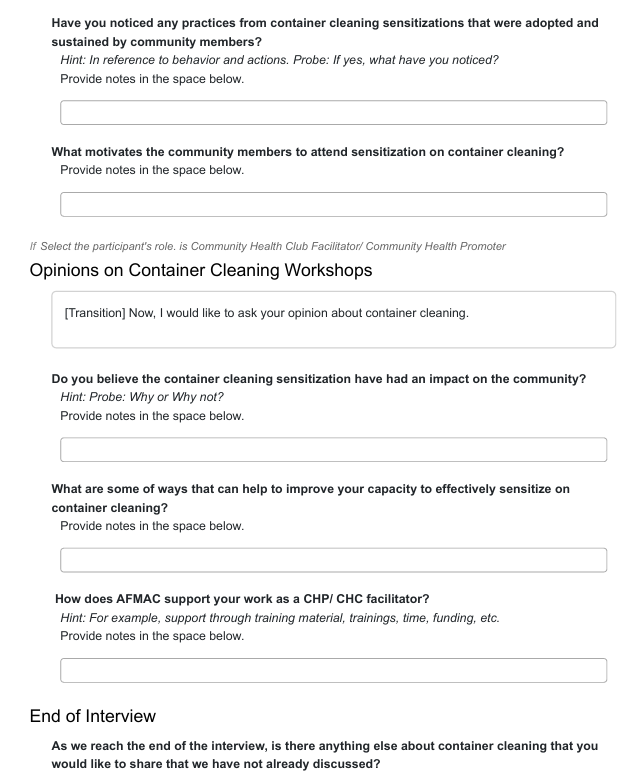

#### Methods S1.4: Household Survey

Container Cleaning Practices HH Survey

Container Cleaning Practices Survey Instructions

Instructions: In this survey, you will be asking households about their current household drinking water storage container cleaning knowledge, attitudes and practices. Please fill out the survey in its entirety. The submit button will indicate the completion of the survey.

Part A: General Information

**Select Interviewer**

Enumerator: "Thank you for accepting to participate in the survey. Before getting started, I need to

record my name, the date, time, and location. Please give me a minute."

◯ Samson Sakala

◯ Esau Banda

**Select Community**

◯ C01

◯ C02

◯ C03

◯ C04

**Select Household**

◯ HH01

◯ HH02

◯ HH03

◯ HH04

◯ HH05

◯ HH06

◯ HH07

◯ HH08

◯ HH09

◯ HH10 ◯ HH11

◯ HH12

◯ HH13

◯ HH14

◯ HH15

◯ HH16

◯ HH17

◯ HH18

◯ HH19

◯ HH20

Part A: General Information

Enumerator: "I will begin by asking you some general questions about your household."

**Respondent's Sex?**

◯ Male

◯ Female

◯ Not Interested

**How old are you?**

◯ 18-24

◯ 25-49

◯ 50-64

◯ 65+

◯ Not Interested

Part A: General Information

**Did you go to school?**

◯ Yes

◯ No

◯ Not Interested

*If Did you go to school? is Yes*

**What is the highest level of education you completed?**

◯ Primary (grade 1-7)

◯ Secondary (grade 8-12)

◯ Tertiary (grade 12 and up)

◯ Not Interested

**Is the female head of house (HoH) able to read and write?**

◯ Yes

◯ No

◯ No female HoH

◯ Not Interested

**Is the male head of house (HoH) able to read and write?**

◯ Yes

◯ No

◯ No male HoH

◯ Not Interested

Part A: General Information

**How many people, including yourself, live in your home?**

Enter a whole number. Enter 7777 for Not Interested.

**Has anyone in your home had diarrhea in the last week?**

◯ Yes

◯ No

◯ I don't know

◯ Not Interested

*If Has anyone in your home had diarrhea in the last week? is Yes*

**How many people have had diarrhea in the last week?**

Enter a whole number. Enter 7777 for Not Interested.

Part B: Household Water Storage Practices

Enumerator: "I would like to ask you questions about the drinking water stored in your household

today."

**Do you collect water in a container that is different from the container you store your drinking water in?**

◯ Yes, they are different

◯ No, they are the same

*If Do you collect water in a container that is different from the container you store your drinking water in? is Yes, they are different*

**Could you show me the container you store your drinking water in?**

◯ Yes

◯ No - END SURVEY

**Why did you choose this storage container?**

☐ Cheap

☐ Convenient

☐ Sturdy

☐ Easy to Use

☐ Other (please specify)

☐ Not Interested

*If Do you collect water in a container that is different from the container you store your drinking water in? (redacted) is*

*No, they are the same or Could you show me the container you store your drinking water in? (redacted) is Yes*

Part B: Household Water Storage Practices

Instructions: In this section, enumerators will make note of observations regarding the household's

water storage container.

**[Observe. In what type of container is the drinking water stored in?]**

◯ Metal pot

◯ Earthen Pot

◯ Jerrycan

◯ Plastic Bottle

◯ Bucket

◯ Barrel

◯ Don't Know

◯ Other (please specify)

*If [Observe. In what type of container is the drinking water stored in?] is one of Don't Know, Other (please specify)*

**[Observe. Is the storage container circular or rectangular?]**

◯ Circular

◯ Rectangular

*If [Observe. Is the storage container circular or rectangular?] (redacted) is Circular*

*or [Observe. In what type of container is the drinking water stored in?] (redacted) is any of Metal pot, Earthen Pot, Plastic Bottle, Barrel, Bucket*

**[Observe. What is the diameter and height of the container?]** *Hint: Check each box to input the values.*

☐ Diameter (cm)

☐ Height (cm)

*If [Observe. In what type of container is the drinking water stored in?] (redacted) is Jerrycan or [Observe. Is the storage container circular or rectangular?] (redacted) is Rectangular*

**[Observe. What is the length, width and height of the container?]** *Hint: Check each box to input the values.*

☐ Length (cm)

☐ Width (cm)

☐ Height (cm)

**[Observe. What is the size of the container opening?]**

◯ very narrow

◯ narrow

◯ wide

◯ very wide

**[Observe. What volume is the container?]**

**[Observe. Is the outside of the container visibly dirty?]**

◯ Yes

◯ No

*If [Observe. In what type of container is the drinking water stored in?] was answered*

Part B: Household Water Storage Practices

**[**

**Observe. Where is the container located?**

**]**

Instructions: In this section, enumerators will make note of observations regarding the household's

water storage container.

◯ Inside, on ground

◯ Inside, raised on table

◯ Outside, on ground

◯ Outside, raised on table

◯ Don't know

◯ Other (please specify)

**[Observe. Is the container cracked?]**

◯ Yes

◯ No

◯ Don't Know

[Observe. On a scale of 1-5, how dirty is the inside of the storage ◯ ◯ ◯ ◯ ◯ container?]	1 2 3 4 5

**[Observe. Note any additional observations about the inside of the water storage container.]** *Hint: Does it have green or black algal growth? Describe the inside of the container.*

*If [Observe. In what type of container is the drinking water stored in?] was answered*

**What is the main source of drinking water used by members of your household?**

◯ Piped into dwelling

◯ Piped to yard/plot

◯ Piped to neighbor

◯ Public tap / stand pipe

◯ Tube well / borehole

◯ Protected dug well

◯ Unprotected dug well

◯ Protected spring

◯ Unprotected spring

◯ Rainwater

◯ Tanker-truck

◯ Cart with small tank

◯ Water kiosk

◯ Surface water (river, dam, lake, pond, stream, canal, irrigation channel)

◯ Bottled water

◯ Sachet water

◯ Large bottle / dispenser refill

◯ Don't know

◯ Other (please specify)

◯ Not Interested

**How long does it take for you or any member of your household to go there, get water, and come back?**

◯ Collects water - Specify how many minutes

◯ Does not collect water

◯ Don't Know

◯ Not Interested

**When was the last time this container was filled/refilled?**

◯ Today

◯ Yesterday

◯ Several days ago

◯ Several weeks ago

◯ Don't Know

◯ Other (please specify)

◯ Not Interested

**Do you or any member of your household clean this container?**

◯ Yes

◯ No

◯ Don't Know

◯ Not Interested

*If Do you or any member of your household clean this container? is one of Yes, Don't Know and [Observe. In what type of container is the drinking water stored in?] was answered*

**Where do you or any member of your household clean this container?**

◯ At home

◯ At the water source

◯ Other (please specify)

◯ Not Interested

**When was the last time you cleaned this container?**

◯ Today

◯ Yesterday

◯ Several days ago

◯ A week ago

◯ Two weeks ago

◯ A month ago

◯ Never

◯ Don't Know

◯ Other (please specify)

◯ Not Interested

*If When was the last time you cleaned this container? isn't Never*

**What do you use to clean this container? [Multiple answers possible]**

☐ Water

☐ Chlorinated Water

☐ Sponge/Brush

☐ Soap

☐ Bleach

☐ Rocks/ Sand

☐ Detergent

☐ Don't Know

☐ Other (please specify)

☐ Not Interested

*If [Observe. In what type of container is the drinking water stored in?] was answered*

**Would you please show me how you normally serve a cup of drinking water?**

◯ Yes

◯ No

*If Would you please show me how you normally serve a cup of drinking water? is Yes*

Instructions: In this section, enumerators will take note of observations regarding the inside of

the water storage container.

**[Observe. how do they get water out of the container?]**

◯ Dips cup into container

◯ Pours water out of container

◯ From container's tap

◯ Other (please specify)

**[Observe. Is there already water present in those containers?]**

◯ Yes

◯ No

**[Observe. How full of water is the container?]**

◯ Empty

◯ Less than half

◯ Approximately half

◯ More than half

◯ Full

◯ Don't Know

**[Observe. Is the inside of the container visibly dirty?]**

◯ Yes

◯ No

*If [Observe. In what type of container is the drinking water stored in?] was answered and Would you please show me how you normally serve a cup of drinking water? is Yes*

**Did someone in your household treat this water in any way?**

◯ Yes

◯ No

◯ Don't know

◯ Not Interested

*If Did someone in your household treat this water in any way? is Yes*

**How was the water treated?**

◯ Boiled

◯ Sand filter ◯ Cloth filter

◯ Ceramic filter

◯ Tablet chlorine ◯ Liquid chlorine

◯ Other filter

◯ Don't know

◯ Other (please specify)

◯ Not Interested

**Approximately how long ago was the water treated?**

◯ Today

◯ Yesterday

◯ Several days ago

◯ Several weeks ago

◯ Don't know

◯ Other (please specify)

◯ Not Interested

*If [Observe. In what type of container is the drinking water stored in?] was answered and Would you please show me how you normally serve a cup of drinking water? is Yes*

In this section, the enumerator will take a photo of the exterior of the water storage container. Then, collect a sample for microbiological analysis from the container. Ensure the sample is collected in the collection bag using sterile technique. Place on ice to return to field lab. Then collect physical and chemical water quality parameters.

**[Take a photo of the exterior of the water storage container.]**

*Hint: The picture should show the sides of the container with cap/lid on.*

☐ Not Applicable

**[Take a photo of the top of the water storage container.]** *Hint: Take photo with the cap/lid off.*

☐ Not Applicable

**[Did you collect the water quality parameters of the drinking water?]**

◯ Yes

◯ No

◯ Other (please specify)

*If [Did you collect the water quality parameters of the drinking water?] is Yes*

Water Quality Parameters

**Record pH**

Enter 7777 for Not Interested.

**Record temperature**

Enter 7777 for Not Interested.

**Record Total Disolved Solids (TDS)**

Select which units you are measuring the TDS. Enter 7777 for Not Interested.

**Electrical Conductivity**

Enter 7777 for Not Interested.

*If [Observe. In what type of container is the drinking water stored in?] was answered*

Part C: Household Water Storage Container Handling Knowledge

Enumerator: " I will ask you questions about your household water container practices."

**Should you clean your drinking water storage containers?**

◯ Yes

◯ No

◯ Don't know

◯ Not Interested

*If Should you clean your drinking water storage containers? is one of Yes, Don't know* **How often should you clean your water storage containers?**

◯ Daily

◯ Weekly

◯ Monthly

◯ Never

◯ Don't know

◯ Not Interested

**Do you think you can get sick from dirty water storage containers?**

◯ Yes

◯ No

◯ Don't know

◯ Not Interested

*If [Observe. In what type of container is the drinking water stored in?] was answered*

*If Do you think you can get sick from dirty water storage containers? is Yes*

**What kind of sickness can you get from drinking water from unclean containers? [Multiple answers possible]**

*Hint: [Probe. Any more? ]*

☐ Diarrhea ☐ Vomiting

☐ Stomach ache

☐ Fever

☐ Cholera

☐ Dehydration

☐ Headache

☐ Influenza

☐ General pain

☐ Parasites

☐ Don't know

☐ Other (please specify)

☐ Not Interested

**If you were to clean your water storage containers, how would you know the container should be cleaned? [Multiple answers possible]** *Hint: [Probe. Any other reason?]*

☐ Water is not clear

☐ Based on time last cleaned

☐ Tastes / smells funny

☐ Outside of container is dirty

☐ Inside of container is dirty

☐ Don't know

☐ Other (please specify)

☐ Not Interested

**How might you know if your water is not safe to drink? [Multiple answers possible]** *Hint: [Probe. Any other reason?]*

☐ Looks dirty

☐ Has bacteria

☐ Bad source

☐ Makes you sick

☐ Not treated

☐ Stored in open container

☐ Tastes / smells bad

☐ Don't know

☐ Other (please specify)

☐ Not Interested

*If [Observe. In what type of container is the drinking water stored in?] was answered*

**Have you ever been taught to clean drinking water storage containers?**

◯ Yes

◯ No

◯ Not Interested

*If Have you ever been taught to clean drinking water storage containers? is Yes*

Container Cleaning Workshop Knowledge

**Where did you learn about how to clean your water storage containers?**

☐ Friends and Family

☐ Workshops

☐ Community meeting

☐ School

☐ Don't know

☐ Other (please specify)

☐ Not Interested

*If Where did you learn about how to clean your water storage containers? does not include Workshops* **Have you ever attended a container cleaning workshop?**

☐ Yes, from Seeds of Hope

☐ Yes, from other organizations

☐ No

☐ Not Interested

*If Have you ever attended a container cleaning workshop? includes any of Yes, from Seeds of Hope or Where did you learn about how to clean your water storage containers? includes all of Workshops*

**How many Seeds of Hope (CHC) workshops have you attended?**

Enter 7777 for Not Interested.

*If Have you ever attended a container cleaning workshop? includes any of Yes, from other organizations or Where did you learn about how to clean your water storage containers? includes all of Workshops*

**How many other workshops have you attended?**

Enter 7777 for Not Interested.

*If Have you ever been taught to clean drinking water storage containers? (redacted) is No or Have you ever attended a container cleaning workshop? includes all of None or How many Seeds of Hope (CHC) workshops have you attended? (magnitude) (redacted) is not blank or How many Seeds of Hope (CHC) workshops have you attended? (magnitude) (redacted) is None is blank*

End of Survey

Enumerator: "We have reached the end of the survey. Thank you very much for your time! We will

now move onto the biofilm sample collection."

**Select to record the end of the survey.**

*If Could you show me the container you store your drinking water in? is No*

End of Survey (No Drinking Water Container Shown)

Enumerator: "We have reached the end of the survey. Thank you very much for your time!"

**Select to record the end of the survey.**

### Glossary

| Biofilm | communities of microorganisms embedded in extracellular polymeric substance (EPS) matrix and are found on the interior surfaces of pipes, taps and water storage containers. Biofilms provide bacteria with a favorable environment against harsh environmental conditions. |
| --- | --- |
| HPC | Heterotrophic Plate Count is an indicator of general microbial content of water including bacteria, yeasts, and molds. |
| HWTS | Household Water Treatment and Safe Storage is an interim intervention focused on point of use that promotes household level water treatment and safe storage to improve drinking water quality and reduce diarrheal diseases. |
| CHC | Community Health Club |
| CHP | Community Health Promoter |
| AFMAC | Africa MANZI Center |
| CAWST | Centre for Affordable Water and Sanitation Technology |
| FGD | Focus Group Discussion |
| KII | Key Informant Interview |

### REFERENCES

(1)	Oliveira, I. M.; Gomes, I. B.; Simões, L. C.; Simões, M. A Review of Research Advances on Disinfection Strategies for Biofilm Control in Drinking Water Distribution Systems. *Water Research* **2024**, *253*, 121273. https://doi.org/10.1016/j.watres.2024.121273.

(2)	Li, Y.; Qu, Y.; Yang, H.; Zhou, X.; Xiao, P.; Shao, T. Combatting Biofilms in Potable Water Systems: A Comprehensive Overview to Ensuring Industrial Water Safety. *Environ Microbiol Rep* **2023**, *15* (6), 445–454. https://doi.org/10.1111/1758-2229.13207.

(3)	Nnaji, C. C.; Nnaji, I. V.; Ekwule, R. O. Storage-Induced Deterioration of Domestic Water Quality. *Journal of Water, Sanitation and Hygiene for Development* **2019**, *9* (2), 329–337. https://doi.org/10.2166/washdev.2019.151.

(4)	Schafer, C. A.; Mihelcic, J. R. Effect of Storage Tank Material and Maintenance on Household Water Quality. *Journal AWWA* **2012**, *104* (9), E521–E529. https://doi.org/10.5942/jawwa.2012.104.0125.

(5)	Manga, M.; Ngobi, T. G.; Okeny, L.; Acheng, P.; Namakula, H.; Kyaterekera, E.; Nansubuga, I.; Kibwami, N. The Effect of Household Storage Tanks/Vessels and User Practices on the Quality of Water: A Systematic Review of Literature. *Environmental Systems Research* **2021**, *10* (1), 18. https://doi.org/10.1186/s40068-021-00221-9.

(6)	String, G.; Domini, M.; Mirindi, P.; Brodsky, H.; Kamal, Y.; Tatro, T.; Johnston, M.; Badr, H.; Lantagne, D. Efficacy of Locally-Available Cleaning Methods in Removing Biofilms from Taps and Surfaces of Household Water Storage Containers. *npj Clean Water* **2020**, *3* (1), 1–11. https://doi.org/10.1038/s41545-020-0061-y.

(7)	Steele, A.; Clarke, B.; Watkins, O. Impact of Jerry Can Disinfection in a Camp Environment – Experiences in an IDP Camp in Northern Uganda. *Journal of Water and Health* **2008**, *6* (4), 559–564. https://doi.org/10.2166/wh.2008.072.

(8)	Meierhofer, R.; Wietlisbach, B.; Matiko, C. Influence of Container Cleanliness, Container Disinfection with Chlorine, and Container Handling on Recontamination of Water Collected from a Water Kiosk in a Kenyan Slum. *Journal of Water and Health* **2019**, *17* (2), 308–317. https://doi.org/10.2166/wh.2019.282.

(9)	String, G. M.; Domini, M.; Badr, H.; Brodsky, H.; Kamal, Y.; Tatro, T.; Johnston, M.; Ogudipe, A.; Nha Vu, T.; K. Wolfe, M.; S. Lantagne, D. Efficacy of Locally-Available Cleaning Methods and Household Chlorination at Inhibiting Biofilm Development in Jerricans Used to Store Household Drinking Water. *Environmental Science: Water Research & Technology* **2021**, *7* (2), 367–383. https://doi.org/10.1039/D0EW00748J.

(10)	Judah, L. A.; Andriambololonirina, C.; Rakotoarisoa, L.; Barrett, L. J. P.; Khaliq, M.; Mihelcic, J. R.; Cunningham, J. A. Occurrence and Mitigation of Bacterial Regrowth in Stored Household Water in Eastern Coastal Madagascar. *Water* **2024**, *16* (11), 1592. https://doi.org/10.3390/w16111592.

(11)	Gärtner, N.; Germann, L.; Wanyama, K.; Ouma, H.; Meierhofer, R. Keeping Water from Kiosks Clean: Strategies for Reducing Recontamination during Transport and Storage in Eastern Uganda. *Water Research X* **2021**, *10*, 100079. https://doi.org/10.1016/j.wroa.2020.100079.

(12)	Stocker, A.; Mosler, H.-J. Contextual and Sociopsychological Factors in Predicting Habitual Cleaning of Water Storage Containers in Rural Benin. *Water Resources Research* **2015**, *51* (4), 2000–2008. https://doi.org/10.1002/2014WR016005.

(13)	Bae, S.; Lyons, C.; Onstad, N. A Culture-Dependent and Metagenomic Approach of Household Drinking Water from the Source to Point of Use in a Developing Country. *Water Research X* **2019**, *2*, 100026. https://doi.org/10.1016/j.wroa.2019.100026.

(14)	Budeli, P.; Moropeng, R. C.; Mpenyana-Monyatsi, L.; Momba, M. N. B. Inhibition of Biofilm Formation on the Surface of Water Storage Containers Using Biosand Zeolite Silver-Impregnated Clay Granular and Silver Impregnated Porous Pot Filtration Systems. *PLoS One* **2018**, *13* (4), e0194715. https://doi.org/10.1371/journal.pone.0194715.

(15)	Peletz, R.; Simunyama, M.; Sarenje, K.; Baisley, K.; Filteau, S.; Kelly, P.; Clasen, T. Assessing Water Filtration and Safe Storage in Households with Young Children of HIV-Positive Mothers: A Randomized, Controlled Trial in Zambia. *PLOS ONE* **2012**, *7* (10), e46548. https://doi.org/10.1371/journal.pone.0046548.

(16)	Peletz, R.; Simuyandi, M.; Simunyama, M.; Sarenje, K.; Kelly, P.; Clasen, T. Follow-Up Study to Assess the Use and Performance of Household Filters in Zambia. *Am J Trop Med Hyg* **2013**, *89* (6), 1190–1194. https://doi.org/10.4269/ajtmh.13-0054.

(17)	CAWST. *Biofilm Infographic: Are you protecting those you serve? | WASH Resources*. WASH Resources. https://washresources.cawst.org/en/resources/f378c5cc/biofilm-infographic (accessed 2025-03-05).

(18)	CAWST. *Safe Drinking Water Storage and Handling | WASH Resources*. WASH Resources. https://washresources.cawst.org/en/module/ce8104f2/safe-drinking-water-storage-and-handling (accessed 2025-03-05).

(19)	Waterkeyn, J.; Cairncross, S. Creating Demand for Sanitation and Hygiene through Community Health Clubs: A Cost-Effective Intervention in Two Districts in Zimbabwe. *Social Science & Medicine* **2005**, *61* (9), 1958–1970. https://doi.org/10.1016/j.socscimed.2005.04.012.

(20)	Brooks, J.; Adams, A.; Bendjemil, S.; Rosenfeld, J. Putting Heads and Hands Together to Change Knowledge and Behaviours: Community Health Clubs in Port-Au-Prince, Haiti. *Waterlines* **2015**, *34* (4), 379–396.

(21)	Acting Officer in Charge of Kaloko Clinic. Top health concerns in Kaloko community., 2024.

(22)	Environmental Health Technician of Mwange Health Post. Top health concerns in Mwange community., 2024.

(23)	Community Heath Assistant of Mushili Health Post. Top health concerns in Chibili community., 2024.

(24)	Sister in-charge of George Clinic. Top health concerns in George community., 2024.

(25)	WASHplus. *Social, Cultural and Behavioral Correlates of Household Water Treatment and Storage*. Sanitation Updates. https://sanitationupdates.wordpress.com/2010/09/16/social-cultural-and-behavioral-correlates-of-household-water-treatment-and-storage/ (accessed 2025-04-08).

(26)	Ntakarutimana, A.; Ekane, N. Performance of Community Health Clubs in Transforming Sanitation and Hygiene Conditions. **2017**.

(27)	Graves, J. M.; Daniell, W. E.; Harris, J. R.; Obure, A. F. X. O.; Quick, R. Enhancing a Safe Water Intervention with Student-Created Visual AIDS to Promote Handwashing Behavior in Kenyan Primary Schools. *Int Q Community Health Educ* **2012**, *32* (4), 307–323. https://doi.org/10.2190/IQ.32.4.d.

(28)	Polly, J. Y.; Nayoan, C. R.; Limbu, R.; Marni, M. Demonstration Method Better Increased Knowledge, Attitude, and Skills on Hand Washing With Soap in Elementary School Students. *Journal of Public Health for Tropical and Coastal Region* **2024**, *7* (3), 249–255. https://doi.org/10.14710/jphtcr.v7i3.22869.

(29)	Niaone, M.; Bendjemil, S.; Rosenfeld, J.; Berggren, R. Community Health Clubs for Water, Sanitation and Hygiene (WASH) Improvement in Rural Burkina Faso. *Annals of Global Health* **2016**, *82* (3). https://doi.org/10.1016/j.aogh.2016.04.352.

(30)	Meierhofer, R.; Rubli, P.; Dreyer, K.; Ouma, H.; Wanyama, K.; Peter-Varbanets, M. Membrane Filtration Reduces Recontamination Risk in Chlorinated Household Water Containers. **2017**.

(31)	Snyder, L. Promoting Hygiene and Sanitation: A Case Study of Community Clubs in Rural Zambia and its Impacts on Child Health Outcomes, The University of North Carolina at Chapel Hill, 2013. https://doi.org/10.17615/nhzd-nc53.

(32)	Hoffmeyer-Zlotnik, J. P. New Sampling Designs and the Quality of Data. *Developments in Applied Statistics/Metodološki zvezki ER* **2003**, *19*.
